# OXA-181 transmission confounded by a stable IncX3 plasmid

**DOI:** 10.64898/2026.08.20.26360670

**Authors:** Tricia S. E. Lee, Liam Nguyen, Brian M. Forde, Toby Maidment, Suifang Ye, Andrew Henderson, E. Geoffrey Playford, Naomi Runnegar, Belinda Henderson, Catherine Watson, Margaret Lindsay, Evan Bursle, Joel Douglas, Jocelyn Hume, David L. Paterson, Timothy Kidd, Bianca Graves, Anna Hume, Michael B Hall, Mark A Schembri, Scott A. Beatson, Patrick N.A. Harris, Leah W. Roberts

## Abstract

OXA-48-like carbapenemases have been historically rare, however steady increases both locally and globally have warranted further investigation into their spread. Here we present the largest genomic analysis of *bla*_OXA-181_-producing bacteria in Australia to date, focusing on a single jurisdiction over seven years (2017 – 2024).

The initial investigation was prompted by an outbreak in 2017, where enhanced genomic surveillance in a single hospital identified 85 outbreak isolates related to an imported *Escherichia coli* ST38, carrying *bla*_OXA-181_ on an IncX3/colKP3 plasmid (previously reported as pOXA181). After four months of intensive infection control, the initial outbreak strain was eliminated. To confirm the outbreak plasmid was also contained, we collected all *bla*_OXA-181_-positive isolates from the same jurisdiction over subsequent years and sequenced with both Illumina and Oxford Nanopore Technologies to investigate clonal and mobile genetic element mediated spread.

While continued surveillance post-2017 did not identify the same *E. coli* strain following the outbreak, pOXA181 plasmids were identified in >70% of surveillance isolates, with minimal genetic changes, which initially suggested local plasmid-mediated spread. Additional comparison to a global collection of pOXA181 plasmids found that epidemiologically unrelated pOXA181 plasmids were near identical, with no rearrangements and low, or no, single nucleotide polymorphisms. This suggests the mutation rate of pOXA-181 is incompatible with recent genomic transmission inference.

This study highlights the current genomic epidemiology and drivers of *bla*_OXA-181_ and further demonstrates the necessity for detailed understanding of plasmid evolutionary rates to inform genomic surveillance.

**Data summary:** All assemblies and raw sequence data has been uploaded under BioProject PRJNA545001 (see accessions in Supplementary Dataset S1).

**Impact Statement:** Here we present one of the largest genomic studies of *bla*_OXA-181_-producing bacteria globally, combining seven years of local genomic surveillance with a global collection of *bla*_OXA-181_-carrying plasmids. We found that the IncX3/colKP3 plasmid carrying *bla*_OXA-181_ is remarkably stable, with epidemiologically unrelated plasmids showing little or no genetic variation. Consequently, plasmids that appear to be recently transmitted based on genomic similarity alone may instead represent independent introductions of a highly conserved plasmid. This finding highlights an important limitation of plasmid-based genomic epidemiology and demonstrates that plasmid sequence similarity cannot necessarily be interpreted as evidence of recent transmission. Our study provides a cautionary example for the increasing use of long-read sequencing and plasmid surveillance in antimicrobial resistance investigations, emphasising the need to consider plasmid evolutionary rates and epidemiological context when interpreting genomic evidence of transmission.

## Introduction

Increasing incidence of antimicrobial resistance (AMR) among Gram-negative bacteria is an issue of global concern^1^. Resistance to carbapenems poses a particularly large threat to public health, as carbapenems remain one of our last-line antibiotics^2^. Three classes of carbapenem-hydrolysing β-lactamases have been described, including class A (such as KPC), class B (also known as metallo-β-lactamases, such as IMP, VIM and NDM), and class D (such as OXA-48-like)^3,4^. OXA-48-like carbapenemases are endemic in certain regions of the world and increasingly detected globally via repeated introduction into non-endemic regions^5,6^. While there are 67 alleles in the OXA-48-like AMR gene family (CARD^7^ as of August 4^th^ 2025), alleles such as OXA-48, OXA-181, and OXA-232 remain the most common.

The *bla*_OXA-181_ gene has been broadly identified across both human and nonhuman samples^6^. OXA-48-like β-lactamases generally hydrolyse penicillins efficiently and carbapenems at relatively low rates, however, hydrolytic activity varies among variants, with OXA-181 exhibiting greater carbapenemase activity than OXA-48 and OXA-232^3,8^. Due to this low-level carbapenemase activity, detection of OXA-48-like enzymes can often be missed by standard phenotypic methods employed routinely in diagnostic laboratories^6,9^. OXA-48-like enzymes are also not susceptible to classical β-lactamase inhibitors, such as clavulanic acid, tazobactam and sulbactam, complicating treatment options if not correctly identified^6,10^.

Reports of OXA-48-like carbapenemases in Australia have been historically rare and generally linked to importation from international travel rather than endemic spread^11,12^. Since the first reported case in 2011, however, cases have been increasing; OXA-48-like detection peaked in 2022 (n=294), with a report in 2024 detecting 255 isolates (AURA^13^, CARAlert^14^), of which at least 35% were *bla*_OXA-181_. This, and the fact that OXA-48-like activity may be going undetected, highlights a growing issue of carbapenem resistance mediated by *bla*_OXA-181_ in Australia.

Here we present a seven-year genomic surveillance study of *bla*_OXA-181_ stemming from an outbreak of *E. coli* carrying both the *bla*_OXA-181_ carbapenemase and *bla*_CTX-M-15_ extended-spectrum beta-lactamase (ESBL) at a major tertiary hospital in Brisbane, Queensland Australia^15^. This study represents the largest genomic characterisation of *bla*_OXA-181_ in Australia to date, providing insights into this carbapenemase and its modes of transmission in healthcare settings.

## Materials and Methods

### Organism identification and antimicrobial susceptibility testing

Full details are provided in the Supplementary Methods (pp.3-4). The OXA-181 outbreak occurred at the Princess Alexandra Hospital (PAH) in Brisbane, Queensland 2017. Rectal swabs were collected and cultured directly on ESBL chromogenic agar. Species identification was performed with VITEK MS (bioMérieux) and antimicrobial susceptibility tests (AST) performed using VITEK 2 GN AST cards (bioMérieux) and interpreted according to EUCAST clinical breakpoints (v8.0). Broth microdilution was performed on the first 6 isolates. Select isolates had a β CARBA test (Bio-Rad), followed by culture on CHROMID CARBA SMART agar (bioMérieux). Isolates positive for either β CARBA or CHROMID underwent an in-house multiplex real-time PCR for key carbapenemase genes^16–19^.

Ongoing screening for carbapenemase-producing Enterobacterales (CPE) was performed with minor changes to the above. From 2024, mSuperCarba (CHROMAgar) replaced CHROMID CARBA SMART and any isolates cultured on mSuperCarba underwent testing using the NG Test Carba-5 lateral flow assay (NG Biotech). All OXA-181 positive isolates collected between June 2017 – Dec 2024 from patients in Queensland were included in this study. MICs were interpreted according to EUCAST guidelines applicable at the time of testing.

### Sequencing, basecalling and quality control

Genomic DNA extraction as well as Illumina and Oxford Nanopore Technologies (ONT) sequencing conditions are described in the Supplementary Methods (pp.4-5). Briefly, genomic DNA for ONT sequencing was extracted with either the MoBio UltraClean Microbial DNA extraction kit or the Roche High Pure PCR Template Preparation Kit. Genomic DNA for Illumina sequencing was extracted using a DSP QIAamp DNA mini kit on a QiaSymphony (Qiagen). Illumina sequencing was performed on an Illumina NextSeq500 (150bp paired end). ONT sequencing on the 2017 samples was performed on a MinION R9.4 flow cell using the SQK-LSK108 kit. Contemporary sequencing on the post-outbreak 2017-2024 samples was done using the SQK-RBK114.96 kit multiplexed on a single PromethION R10.4.1 flow cell.

### Illumina analysis

All raw Illumina reads were checked for quality using FastQC (v0.11.5)^20^. Reads were trimmed using Nesoni v0.133^21^. Trimmed reads were *de novo* assembled using Spades v3.11.1^22^. *S*equence typing was performed using the tool mlst v2.1^23^ ^24^. Plasmid incompatibility (Inc) types, resistance genes and virulence genes were determined using Abricate v0.8^25^ with the following databases: Plasmidfinder^26^, VFDB^27^, Resfinder^28^ and ARG-ANNOT^29^ (updated April 2018). O and H types were determined by screening the EcOH database (updated 18 January 2019) using Abricate. K type was determined using Kaptive (v0.5.1)^30^. *fimH* antigen typing was determined using Fimtyper (v1.1)^31^. Core single nucleotide polymorphisms (SNPs) were called using Nesoni against the *de novo* assembly from PAH outbreak isolate MS14445 from a core genome of 4,651,241 base pairs (bps).

### Nanopore analysis

Nanopore analysis during the 2017 outbreak was performed as per the gold standard approach at the time and is described in Supplementary Methods (p.5). Nanopore reads from the post-outbreak dataset were basecalled using Dorado (v1.1.1) using the super-accuracy model (dna_r10.4.1_e8.2_400bps_sup@v5.2.0). Reads were checked for quality using Nanoq v0.10.0^32^ and taxonomically profiled with sylph v0.8.1^33^ (using the v0.3-c1000-gtdb-r214.syldb database) before being filtered using Seqkit^34^ (v2.10.0) to remove reads ≤500bp. Trimmed reads were *de novo* assembled with Autocycler v0.5.1^35^. Contigs containing *bla*_OXA-181_ was determined using AMRFinderPlus v4.0.23^36^. *De novo* assemblies were quality checked with QUAST v5.3.0^37^ and checkM2 v1.1.0^38^. Plasmids were screened for replicon type using Plasmidfinder^26^ (v2.1.6) whilst the chromosomal contigs were sequence typed using mlst^23^ (v2.23.0).

### AllTheBacteria screening

Screening public data for closely related *E. coli* to the outbreak strain is described in the Supplementary Methods (p.6). Briefly, the PAH isolate MS14441 chromosome was compared to all *E. coli* from AllTheBacteria (ATB; release 0.2; n=315,066)^39^ using Skani (v0.3.0)^40^. Parsnp (v2.1.5)^41^, Gubbins (v3.4.3)^42^ and RAxML (v8.2.12)^43^ were used to build a phylogeny of the PAH isolate MS14441 and 207 closely related public *E. coli* genomes from ATB.

### Plasmid analysis

Lexicmap^44^ (v0.7.0) was used to search the *bla*_OXA-181_ gene against PLSDB^45^ (v2; 2024_05_31_v2; n=72,360 plasmids), filtering for 100% coverage and 100% identity. Plasmid networks were generated using Pling^46^ (v2.0.0). Snippy (v4.6.0) (https://github.com/tseemann/snippy) was run using the snippy-multi option with pMS14441 as the reference against all post-outbreak IncX3/colKP3 plasmid assemblies. Snp-dists (v1.2.0) (https://github.com/tseemann/snp-dists) was used on the Snippy core full alignment to create a distance matrix, which was then used to generate the clustermap using python seaborn^47^ plotting.

## Results

### Preliminary detection of OXA-181 via unusual antibiogram

Over a single week in May 2017, eight patients in a Brisbane tertiary referral hospital (Princess Alexandra Hospital; PAH) were identified by routine screening as newly colonised with ESBL-producing *E. coli* displaying an unusual and identical antibiogram (Table 1; Supplementary Results p.7). Whole genome sequencing found all *E. coli* to be ST38 (phylogroup D) and within 5 single nucleotide polymorphisms (SNPs), suggesting direct transmission (Supplementary Figure 1). All carried the ESBL gene *bla*_CTX-M-15_, the OXA-48-like carbapenemase *bla*_OXA-181_ and the plasmid-mediated quinolone resistance gene *qnrS1,* in addition to IncX3, colKP3 and IncI1 type plasmids (Table 2). O-antigen, capsule and H typing identified the strain as O153var1:K14:H9. Fim typing identified the FimH54 adhesin.

**Table 1:** Broth microdilution MICs for the initial *E. coli* isolates.

| Antimicrobial | MS14441<br>MS14443<br>MS14445<br>MS14446<br>MS14447 | MS14442 |
| --- | --- | --- |
|  | MIC<br>(µg/mL) |  |
| Amikacin | 4 | 4 |
| Gentamicin | 1 | 1 |
| Tobramycin | 1 | 1 |
| Ciprofloxacin | 0.5 | 0.5 |
| Levofloxacin | 1 | 1 |
| Piperacillin/<br>tazobactam<sup>a</sup> | 64 | 64 |
| Amoxycillin/<br>Clavulanic acid<sup>b</sup> | 256 | 256 |
| Cefepime | 16 | 16 |
| Ceftazidime | ≥ 32 | ≥ 32 |
| Cefotaxime | ≥ 16 | ≥ 16 |
| Ceftolozane/<br>tazobactam<sup>a</sup> | 4 | 4 |
| Meropenem | 0.5 | ≥ 32<sup>c</sup> |
| Doripenem | 0.25 | ≥ 16<sup>c</sup> |
| Imipenem | 0.25 | ≥ 32<sup>c</sup> |
| Ertapenem | 2 | ≥ 8<sup>c</sup> |
| Aztreonam | ≥ 32 | ≥ 32 |
| Tigecycline | 2 | 2 |
| Colistin | 0.12 | 0.12 |
| Sulfamethoxazole/<br>trimethoprim | 0.12 | 0.12 |

<sup>a</sup>Tazobactam concentration fixed at 4 µg/ml; <sup>b</sup>Clavulanic acid concentration fixed at 2 µg/ml; <sup>c</sup>see Supplementary Results (p.7) regarding MS14442 meropenem resistance mechanisms

**Table 2:** Initial 6 isolates sequenced in response to possible outbreak.

| Strain | MS14441 | MS14442<sup>1</sup> | MS14443 | MS14445 | MS14446 | MS14447 |
| --- | --- | --- | --- | --- | --- | --- |
| Date | May 2017 | May 2017 | May 2017 | May 2017 | May 2017 | May 2017 |
| Location | Hospital C | Hospital B | PAH/W5D | PAH/W5D | PAH/W5D | PAH/W5D |
| Source | Rectal swab |  |  |  |  |  |
| Sequence type | 38 |  |  |  |  |  |
| O-antigen | O153var1* |  |  |  |  |  |
| Capsule (K) | K14<sup>+</sup> |  |  |  |  |  |
| Flagellin (H) | H9* |  |  |  |  |  |
| fimH antigen | fimH54 |  |  |  |  |  |
| *bla*<sub>CTX-M-15</sub> | + | + | + | + | + | + |
| *bla*<sub>OXA-181</sub> | + | + | + | + | + | + |
| *IncX3* | + | + | + | + | + | + |
| *colKP3* | + | + | + | + | + | + |
| *Incl1* | + | + | + | + | + | + |
| SNP Distances<sup>2</sup> | REF | 4 | 0 | 1 | 0 | 0 |

<sup>1</sup>Strain MS14442 had MICs of $\geq 32$ , $\geq 16$ , $\geq 8$ and $\geq 32$ for meropenem, doripenem, ertapenem and imipenem respectively. <sup>2</sup>SNP distances relative to reference (REF) MS14441. Predicted consequences of SNPs provided in supplementary Table 1. \*100% coverage, >98% nucleotide identity. + 68% coverage, 94% nucleotide identity.

### OXA-181 carried on an IncX3/colKP3 plasmid pOXA181

Three isolates (named MS14441-3) were sequenced on an ONT MinION to determine the plasmid profiles and genomic contexts of the resistance genes. All three isolates contained a ∼51.4 kb IncX3/colKP3 plasmid carrying both *bla*_OXA-181_ and *qnrS1* in a Tn*6361*-like transposon^48^, with 99.96% average nucleotide identity (ANI) compared to pOXA181 (NZ_KP400525), originally isolated in China^49^. Conversely, the location of *bla*_CTX-M-15_ was chromosomal, with a complete IS*Ecp1* insertion upstream and in proximity to a second *qnrS1* gene. All isolates also carried a ∼106 kb IncI1 plasmid that did not carry any genes known to be of clinical significance but may have been responsible for the introduction of *bla*_CTX-M-15_ into the cell (see Supplementary Results p.8). Analysis of the ∼4.9 Mbp chromosomes of MS14441-3 found that they shared 99% nucleotide identity and 100% coverage with exception to MS14443 (99% coverage). All MS14441-3 contained a virulence factor profile consistent with extra-intestinal pathogenic *E. coli* (ExPEC) (see Supplementary Results p.8 and Supplementary Figure 2).

### Hospital-wide screening determines widespread transmission of *E. coli* ST38

Confirmation of an outbreak by WGS prompted increased infection prevention and control interventions including intensive screening of the entire hospital in June 2017, from both patients and environmental sources (Figure 1). WGS of 110 screening isolates identified 84 as *E. coli* ST38. Of these, 78 (93%) were related to the outbreak, corresponding to 72 patients and two isolates obtained from bathroom environments. Overall, nineteen wards in five different buildings at the PAH, four other public hospitals, and two non-Queensland Health Hospitals in South-East Queensland were involved. All patients at hospitals other than PAH had been recently transferred from PAH (Figure 1).

**Figure 1:**
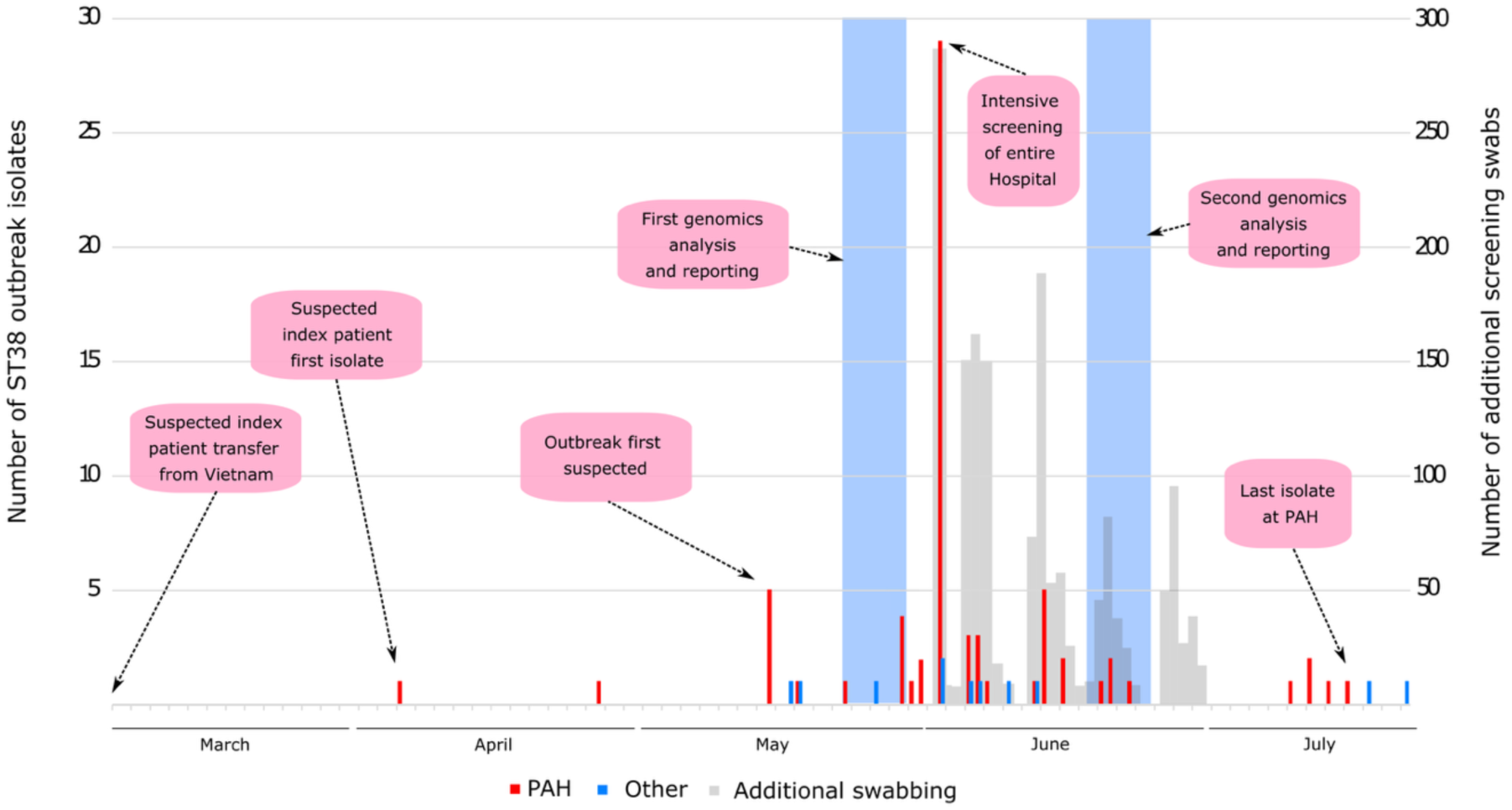
Timeline of ST38 OXA-181 ESBL *Escherichia coli* outbreak: The outbreak is predicted to have lasted from March to July of 2017 but was not detected until mid-May. Isolate numbers are given by the coloured bars (PAH = solid red, other hospital = solid blue) based on the left-hand y-axis. The number of additional swabs are displayed as grey bars based on the right-hand y-axis. The transparent blue bars represent timepoints when genomic analyses was undertaken.

Of 84 *E. coli* ST38 isolates with WGS performed, 82 had a maximum core SNP difference of 7 SNPs to the nearest isolate, with a majority of the isolates identical at the core genome level (based on a core genome of 4,651,241 bp) (Figure 2, Supplementary Figure 3 and Supplementary Dataset S1). Overall, the majority of isolates had a consistent plasmid and AMR gene profile (pOXA181 n=82/84, IncI1 n=84/84, *bla*_OXA-181_ n=80/84, *bla*_CTX-M-15_ n=84/84 and *qnrS1* n=84/84) (Supplementary Figures 4 and 5). Additional results on plasmid and phage differences are described in the Supplementary Results (pp.8-9).

**Figure 2:**
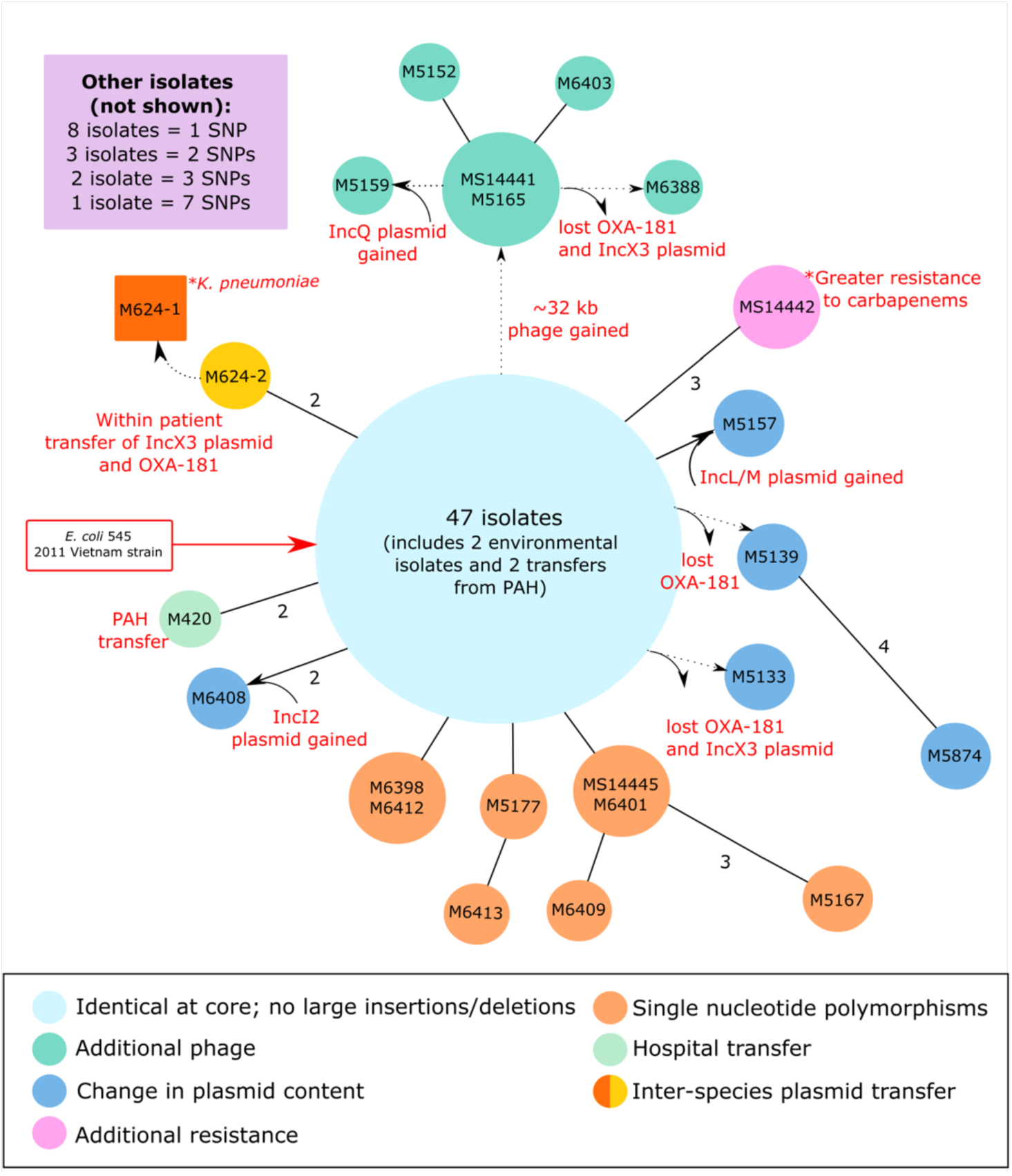
Overall outbreak relationship matrix for ST38 *E. coli*: Isolate reads were mapped to an ST38 reference *E. coli* from the outbreak (MS14445) in order to determine core SNP differences. Circles represent isolates, with the size of the circle broadly equivalent to the number of isolates. Solid lines denote 1 core SNPs difference, except where specified. Dotted lines denote other genomic changes (such as acquisition of a phage or plasmid) as indicated. Due to space constraints remaining isolates are summarised in the top-left corner box.

The final outbreak isolate was detected at a non-Queensland Health hospital in July 2017. Patient clinical outcomes are described in the Supplementary Results (p.9). Ongoing genomic surveillance for all new ESBL-producers and CPE following the outbreak revealed no ongoing transmission of this ST38 clone.

### Presumed international source for ST38 strain carrying OXA-181

To identify the source of the outbreak strain, detailed epidemiological and genomic investigations were conducted (Supplementary Results p.9). Briefly, matching of historical antibiograms identified a patient transferred from Vietnam who, in early April 2017, was found to carry an *E. coli* isolate with an identical antibiogram to the outbreak strain. While no samples were available for testing, comparison of the MS14441 chromosome to the NCBI non-redundant nucleotide database identified a match to the *E. coli* strain Ecol_545 (GenBank CP018976.1), isolated from a patient in Vietnam in 2011, which shared 99.93% average nucleotide identity (ANI) over >95% of the MS14441 chromosome. Further screening of the AllTheBacteria database (v0.2) identified a close match to another isolate from Vietnam (SAMEA5377147; 99.96% ANI, 97.04% alignment fraction), supporting the hypothesis that this strain may have been introduced to the hospital by the putative index patient who had been recently hospitalised in Vietnam.

### Ongoing surveillance reveals steady OXA-181 persistence in Queensland

We continued surveillance for *bla*_OXA-181_ beyond the original outbreak to identify plausible plasmid spread. Between 2017-2024, all *bla*_OXA-181_ positive isolates (n=30) were collected and sequenced on an ONT PromethION flow cell (hereafter referred to as the post-outbreak dataset; Supplementary Figure 10). Specific details of the post-outbreak dataset, including species and MIC, are provided in the Supplementary Results (pp.11-12).

In the post-outbreak dataset, the *bla*_OXA-181_ gene was identified mainly on plasmids (n=27/30, Supplementary Results p.12). Most plasmids carrying *bla*_OXA-181_ were the same incompatibility type as the outbreak plasmid (IncX3/colKP3, n=21), with variant plasmids including IncX3/colKP3/IncR/IncFIA (n=1), IncX3 (n=1), colKP3 (n=1), IncFIB/colKP3 (n=1), IncFII/colKP3 (n=1), and IncAC_2/colKP3 (n=2). A single sample (pQ181_19) contained *bla*_OXA-181_ on two different plasmids (IncX3/colKP3, IncFIB/colKP3). Analysis of the genetic relatedness of the 28 *bla*_OXA-181_ plasmids with the outbreak plasmid using Pling, at a threshold of four rearrangements, identified four subcommunities, the largest containing all IncX3/colKP3 plasmids clustered together with the original outbreak plasmid (n=24), suggesting common ancestry (Figure 3). *bla*_CTX-M-15_ was found in 46% (n=14) of the post-outbreak isolates with 57% (n=8) located on the chromosome, 35% (n=5) on a plasmid separate to the plasmid containing *bla*_OXA-181_, and a single isolate (n=1) with *bla*_CTX-M-15_ on the same plasmid containing *bla*_OXA-181_. This plasmid (pQ181_28) carrying both *bla*_CTX-M-15_ and *bla*_OXA-181_ appeared twice the size of our regular IncX3/colKP3 plasmid with two extra replicons (IncR, IncFIA), suggesting a fusion plasmid (see Supplementary Results p.13; Supplementary Figure 12). The *bla*_OXA-181_ gene was found in a Tn*6361*-like transposon on plasmids in our dataset, irrespective of its broader genetic context (n=28) (Supplementary Results p.13).

**Figure 3:**
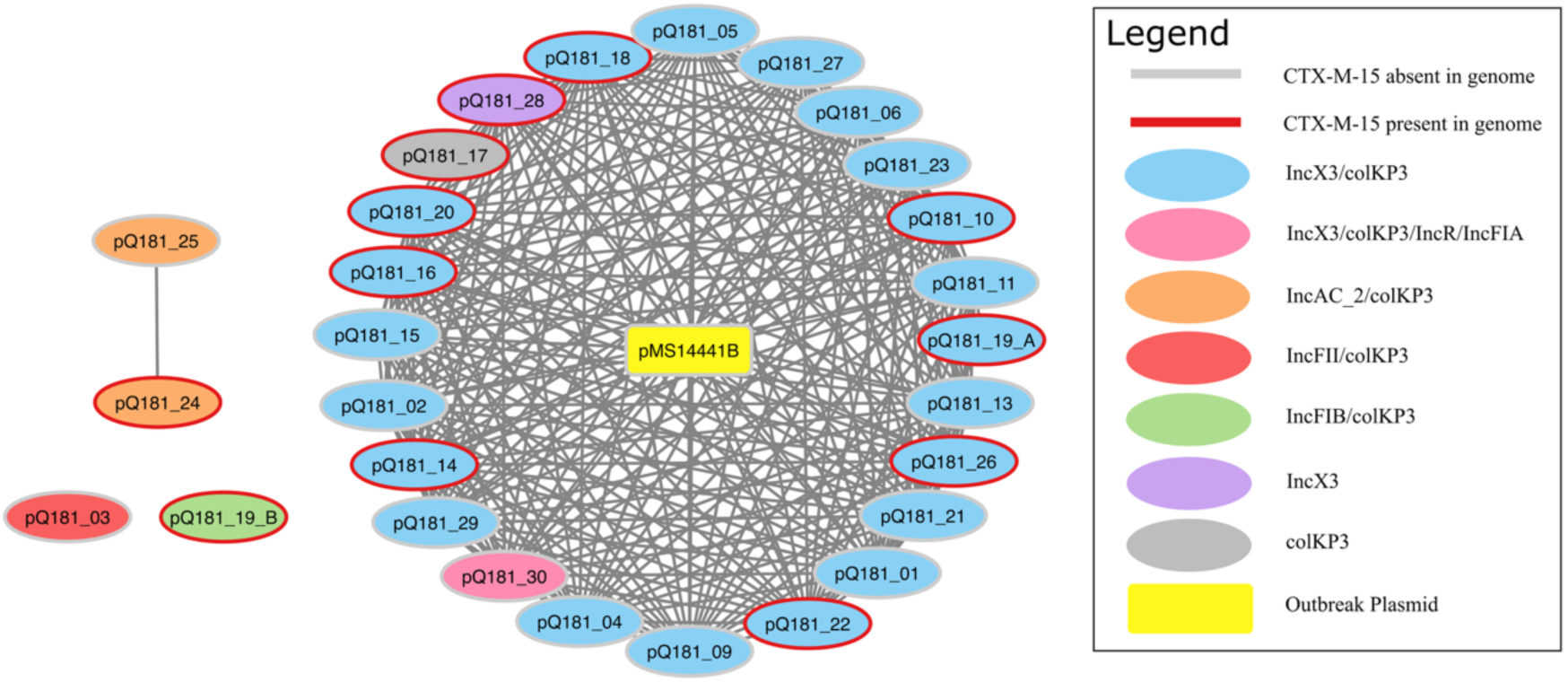
Pling network of OXA-181-carrying plasmids from the post-outbreak dataset. Nodes are colour-coded for each plasmid replicon type. Plasmids whose chromosome/plasmid contained *bla*_CTX-M-15_ had their nodes outlined in red. The outbreak plasmid is highlighted in yellow and sits in the biggest cluster (n=24).

To determine if the post-outbreak IncX3/colKP3 plasmids were the result of continued spread of the outbreak plasmid versus independent introductions, we contextualised our dataset with all IncX3/colKP3 plasmids carrying *bla*_OXA-181_ from PLSDB (n=114). We used Pling with a threshold of zero rearrangements on the combined set of both our plasmids (n=24 related to the PAH outbreak) and 114 global plasmids (Supplementary Figure 13). This identified 36 subcommunities, with 98 plasmids (71%) in a single large subcommunity, including 12 plasmids from our post-outbreak dataset and the original outbreak plasmid. The global plasmids in this subcommunity (n=85) were collected from 22 countries spanning 2017-2024. Using Snippy to identify SNP clusters within this subcommunity revealed that approximately half (n=66) of the samples formed zero-SNP clusters, including a large zero-SNP cluster (n=60) containing plasmids from our dataset (n=5) and publicly available plasmids (n=55) (Figure 4). All post-outbreak plasmids possessed less than five SNPs to their next nearest plasmid in the subcommunity. There was no clear clustering according to bacterial host species, geographical location or collection year.

**Figure 4:**
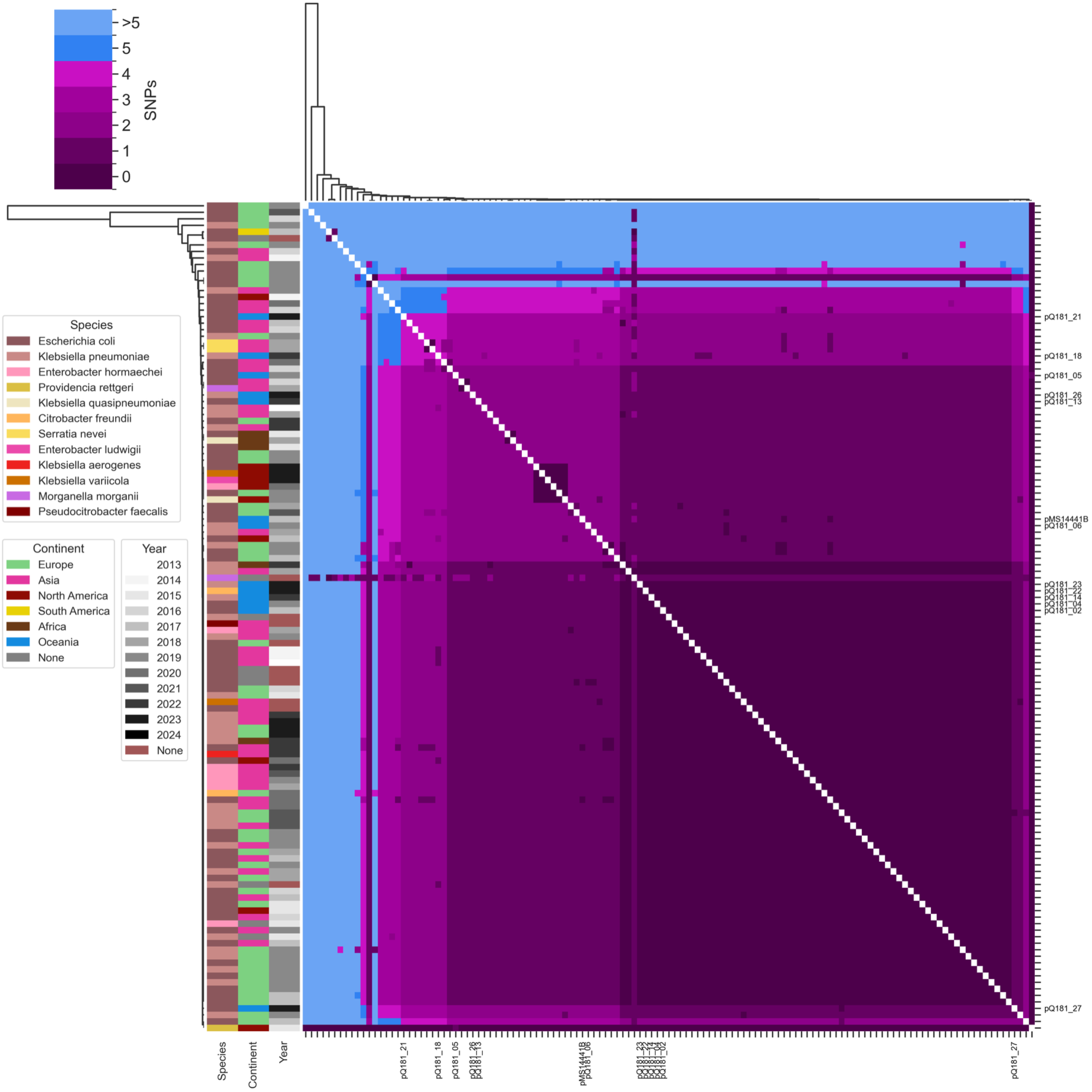
Clustermap representing SNP distances between post-outbreak plasmids, the outbreak plasmid (pMS14441B), and global pOXA-181 plasmids from PLSDB. Only the post-outbreak plasmids and outbreak plasmid are labelled along the axes. Bacterial host species, continent of collection, and the year of collection are colour-coded on the left as per their respective legends. The colour bar at the top indicates the range of pairwise SNP distances between plasmids – from 0 to >5.

The remaining 12 post-outbreak plasmids that were not present in this primary large subcommunity (n=98) were investigated and found to cluster in 10 other subcommunities, where they were mostly standalone members (See Supplementary Results p.13).

## Discussion

Resistance to carbapenems is a major public health threat, particularly in healthcare settings. Here we explored seven years of *bla*_OXA-181_ in Queensland, Australia, following a hospital outbreak in 2017. The original outbreak strain evaded initial detection given its relatively low-level carbapenem resistance and thus spread widely before the outbreak was detected. The identification of *bla*_OXA-181_ on an IncX3/colKP3 plasmid and its potential to spread undetected prompted our additional exploration of modern strains given the increasing prevalence of this gene in Australia.

Prior to this outbreak, the incidence of *E. coli* ST38 or *bla*_OXA-181_ was relatively rare in Australia. The earliest cases of *bla*_OXA-181_ date back to a single reported case in Queensland in 2013^12^ and international transfer of *bla*_OXA-181_ in New Zealand in 2011^50^. It is now one of the most prevalent blaOXA-48-like carbapenemases in Australia, accounting for >50% of OXA-48-like CPE from sequenced isolates in 2024 (CARAlerts^14^). Despite its growing prevalence, it is rarely reported with *E. coli* ST38, which is more frequently reported carrying the ESBL gene *bla*_CTX-M_ and a chromosomally inserted *bla* ^51–53^. *E. coli* ST38 is a globally dispersed lineage that has been consistently reported in Australia since the early 2000s, albeit at low levels from human and non-human sources^54–57^. Reported cases of both ST38 and *bla*_OXA-181_ globally and in Australia, however, are incredibly rare; we could only find a single mention of *bla*_OXA-181_ ST38 *E. coli* in prior literature, which revealed two isolates from a study in broiler chickens^58^. Whether this is due to an under-reporting of ST38 *bla*_OXA-181_ (undetected or under sampled) or a true low prevalence is unknown.

The source of this outbreak was likely a patient transferred from Vietnam in the two months prior to detection, and not from a community source. The concordance of both the epidemiological investigation (into isolates in the preceding months with similar antibiograms) as well as the genomics investigation (into publicly available genomic data) supports this conclusion. Chowdhury et al (2023)^56^ also later conducted a global phylogenetic analysis of ST38 *E. coli*, identifying two highly similar strains of Vietnamese origin that differed by <20 SNPs from our outbreak strain. However, we cannot ignore the global distribution of the ST38 *E. coli* lineage, and its sporadic uptake of the pOXA-181 plasmid. Without further evidence, this result will remain highly probable but ultimately unconfirmed.

Advances in the quality and affordability of long-read sequencing technologies have made plasmid surveillance a reality in public health settings. The continued detection of *bla*_OXA-181_ in the absence of the ST38 outbreak strain suggested a plasmid source of ongoing transmission. Indeed, our pOXA-181 plasmids were highly identical; the majority contained zero rearrangements and zero SNPs, which is usually an indication of transmission or recent ancestry. However, when tested against other publicly available pOXA-181 plasmids, which we assume have no immediate epidemiological connection, we also found little to no genetic changes. This instead reveals that pOXA-181 is a highly stable plasmid and, as such, transmission cannot be reliably determined from sequencing data alone. This is similar to that observed for pOXA-48, a highly stable IncL plasmid, where transmission has been inferred using rare SNP variants in core regions^59^. In our study, we could not identify any genetic variants that could indicate transmission directionality. Based on this analysis alone, we cannot confirm whether the pOXA-181 plasmid has seeded the environment of the PAH, or whether pOXA-181 is being continually imported from external sites, or a combination of both.

The discovery of a single plasmid containing both *bla*_OXA-181_ and *bla*_CTX-M-15_ has only been reported once previously^60^. We show that our plasmid containing *bla*_OXA-181_ and *bla*_CTX-M-15_ is a unique fusion of a pOXA-181 plasmid with an IncR/IncFIA. As the combination of these two resistance genes has been suggested to result in stronger resistance to carbapenems^61^, the emergence of fusion plasmids (especially those with a broad host range) could aid in dissemination of resistance. However, the rarity in which they have emerged thus far suggests that there may be selection against plasmids co-carrying these genes. A core limitation of the post-outbreak dataset was that two-thirds of the isolates had not undergone E-tests and three isolates were not tested with either disc diffusion testing or E-tests. This restricted our ability to make broader inferences about the effect of CTX-M-15/OXA-181 co-occurrence, or measure MIC differences in isolates carrying multiple copies of either OXA-181 and/or CTX-M-15.

Ultimately, this study emphasizes the need to maintain both clonal and plasmid surveillance. With the close cooperation of the diagnostic microbiology laboratory, hospital executives, clinicians, infectious diseases and infection control teams, as well as close networks between other local diagnostic microbiology labs and infection control services, WGS contributed to the rapid containment of the outbreak, to the extent that the ST38 clone has not been detected since. Finally, plasmid surveillance is a critical addition to outbreak surveillance but needs to be interpreted cautiously and with sufficient genomic context. Further work is needed to establish best practices for tracking plasmids in healthcare settings.

## Data Availability

All assemblies and raw sequence data has been uploaded under BioProject PRJNA545001.

## Ethical approval

Ethics approval with waiver of consent was provided by the Human Research Ethics Committee of the Royal Brisbane & Women’s Hospital (HREC/16/QRBW/253). Surveillance beyond the PAH outbreak was performed by Pathology Queensland under the Queensland Genomics Surveillance Program (HREC/17/QFSS/6), with additional sample collection approval from The Queensland Children’s Health Human Research Ethics Committee (HREC/2022/QCHQ/85249).

## Author Contributions

T.S.E.L: Data curation, Formal analysis, Visualization, Investigation, Writing – original draft, Writing – review & editing; L.N: Data curation, Formal analysis, Writing – review & editing; B.M.F: Data curation, Validation, Supervision, Writing – review & editing; T.M: Investigation, Writing – review & editing; S.Y: Data curation, Investigation, Writing – review & editing; A.H. Project administration, Writing – review & editing; E.G.P: Project administration, Methodology, Writing – review & editing; N.R: Data curation, Investigation, Resources, Project administration, Methodology, Writing – review & editing; B.H: Project administration, Methodology, Writing – review & editing; C.W: Project administration, Methodology, Writing – review & editing; M.L: Project administration, Methodology, Writing – review & editing ; E.B: Project administration, Methodology, Writing – review & editing; J.D: Data curation, Investigation, Resources, Project administration, Methodology, Writing – review & editing; J.H: Investigation, Writing – review & editing; D.L.P: Funding acquisition, Project administration, Resources, Writing – review & editing; T.K: Data curation, Investigation, Writing – review & editing; B.G: Data curation, Investigation, Writing – review & editing; A.H: Data curation, Investigation, Writing – review & editing; M.B.H: Conceptualization, Data curation, Formal analysis, Methodology, Software, Supervision, Writing – review & editing; M.A.S: Conceptualization, Funding acquisition, Project administration, Resources, Supervision, Writing – review & editing; S.A.B: Conceptualization, Funding acquisition, Project administration, Resources, Supervision, Writing – review & editing; P.N.A.H: Conceptualization, Funding acquisition, Project administration, Resources, Supervision, Writing – review & editing; L.W.R: Conceptualization, Data curation, Formal analysis, Funding acquisition, Investigation, Methodology, Project administration, Resources, Supervision, Validation, Visualization, Writing – original draft, Writing – review & editing.

## Acknowledgements

We are highly indebted to the tireless commitment and flexibility of the entire PA infection control team and PA Microbiology Laboratory, who were pivotal in controlling and terminating the outbreak. This publication made use of the PubMLST website (https://pubmlst.org/) developed by Keith Jolley and sited at the University of Oxford. The development of that website was funded by the Wellcome Trust. This work was supported by resources provided by the University of Queensland Research Computing Centre’s Bunya supercomputer, with funding from the University of Queensland, Brisbane, Australia^62^.

