## Supplementary Materials for "OXA-181 transmission confounded by a stable IncX3 plasmid"

Lee et al.

**Supplementary Methods:** 3
*Organism identification and antimicrobial susceptibility testing:* 3
*Broth microdilutions:* 4
*Genomic DNA extraction:* 4
2017 outbreak: 4
2017-2024 post-outbreak: 4
*Illumina sequencing:* 5
2017 outbreak: 5
2017-2024 post-outbreak: 5
*Nanopore MinION sequencing:* 5
2017 outbreak: 5
2017-2024 post-outbreak: 5
*Nanopore analysis during 2017 outbreak:* 5
*Screening public data (AllTheBacteria) for closely related E. coli:* 6

**Supplementary Results:** 7
*Initial detection of ESBL and carbapenemase-producing E. coli* 7
*Nanopore sequencing to fully characterize three representative ST38 isolates* 8
*The IncI1 plasmid may have introduced blaCTX-M-15 into the cell* 8
*Additional genetic differences in outbreak dataset:* 8
SNPs and structural variants in SS17M6399 and SS17M6415 8
Acquisition of different plasmids in three isolates 9
*Clinical outcomes* 9
*International source of outbreak isolate* 9
*No clonal transmission of post-outbreak dataset* 11
*Repeat samples from patients in post-outbreak dataset* 11
*Post-outbreak dataset MIC results* 12
*OXA-181 on the chromosome* 12
*Fusion plasmid carrying both $bla_{OXA-181}$ and $bla_{CTX-M-15}$* 13
*$bla_{OXA-181}$ is carried in a truncated variant of Tn6361* 13

**Supplementary Tables:** 15
*Supplementary Table 1: Predicted consequences of SNPs in Supplementary Figure 1* 15
*Supplementary Table 2: PHASTER comparison of phage sequences in SS17M6399 compared with draft Nanopore reference strain MS14441* 16
*Supplementary Table 3: SNP consequences in blood isolate M94949* 17

**Supplementary Figures:** 18
*Supplementary Figure 1: Relationship matrix of initial 6 E. coli isolates* 18
*Supplementary Figure 2: Comparison of entire chromosomes of K-12 MG1655, MS14441, MS14442 and MS14443* 19

*Supplementary Figure 3: BRIG comparison of all ST38 outbreak isolates to MS14441 chromosome* 20
*Supplementary Figure 4: BRIG comparison of all ST38 outbreak isolates to pMS14441A (Incl1 plasmid)* 21
*Supplementary Figure 5: BRIG comparison of all ST38 outbreak isolates to pMS14441B (IncX3 plasmid)* 22
*Supplementary Figure 6: BRIG comparison of SS17M5159 draft assembly to pO111-CRL-115* 23
*Supplementary Figure 7: Context of antibiotic resistance genes in SS17M5159 based on reference plasmid pO111-CRL-115* 24
*Supplementary Figure 8: Contextualizing ST38 outbreak isolates against publicly available E. coli complete genomes* 25
*Supplementary Figure 9: RAxML tree of 207 genomes from AllTheBacteria* 26
*Supplementary Figure 10: timeline of post-outbreak dataset* 27
*Supplementary Figure 12: Fusion plasmid containing $bla_{OXA-181}$ and $bla_{CTX-M-15}$* 29
*Supplementary Figure 13: Structural comparison of post-outbreak plasmids with the blaOXA-181 carrying transposon Tn6361.* 30
References: 32

#### Supplementary Methods:

##### Organism identification and antimicrobial susceptibility testing:

The OXA-181 outbreak occurred at the Princess Alexandra Hospital (PAH) in Brisbane, Queensland 2017. Rectal swabs were collected between May to September 2017 and cultured directly on ESBL chromogenic agar for 24 hours in $O_2$ at 35$^{o}$C (bioMérieux). Species identification was performed with VITEK MS (bioMérieux) and antimicrobial susceptibility tests (AST) performed using VITEK 2 GN AST cards (bioMérieux) and interpreted according to EUCAST clinical breakpoints (v8.0). Broth microdilution was performed on the first 6 isolates using a custom made Sensititre plates (Thermo Fisher) (see Supplementary Methods). Isolates with a VITEK 2 meropenem minimum inhibitory concentration (MIC) >0.125 mg/L had a β CARBA test (Bio-Rad), followed by culture on CHROMID CARBA SMART agar (bioMérieux) in $O_2$ at 35$^{o}$C for 18-24 hours. Isolates positive for either β CARBA or CHROMID underwent an in-house multiplex real-time PCR for OXA-48-like, KPC, NDM, IMP-4, and VIM genes[10–13]. OXA-48-like *E. coli* isolates and all ESBL-positive *E. coli* (and clinically relevant co-colonizing species) from rectal swabs between 2$^{nd}$ to 30$^{th}$ June 2017 were sent for whole genome sequencing (WGS).

Ongoing screening for carbapenemase-producing Enterobacterales (CPE) was performed at Pathology Queensland with minor changes to the methods described above. Any isolates cultured on ESBL chromogenic agar were subsequently cultured on selective chromogenic CPE agar regardless of the Meropenem MIC. From 2024, mSuperCarba (CHROMAgar) replaced CHROMID CARBA SMART and any isolates cultured on mSuperCarba underwent testing for the presence of OXA-48-like, KPC, NDM, IMP-4, and VIM enzymes using the NG Test Carba-5 lateral flow assay (NG Biotech). All suspected CPE identified from laboratories across QLD were referred to the Central microbiology laboratory for molecular or WGS confirmation. All OXA-181 positive isolates collected between June 2017 – Dec 2024 from patients in Queensland were included in this study. MICs were interpreted according to EUCAST guidelines applicable at the time of testing.

Overall, while the majority of isolates were obtained from screening samples (n=102), a small number of clinical samples were also included (blood culture n=6, sputum n=2, urine n=9, wound n=1) in addition to two environmental samples (Supplementary Dataset S1).

#### Broth microdilutions:

Isolates were subbed from beads stored at -80$^{o}$C onto Horse Blood Agar (HBA, BioMerieux). Following incubation at 35$^{o}$C $O_2$ for 24 hours, single colonies were then inoculated into sterile saline and adjusted to a Macfarland concentration of 0.5. A final concentration of 1 x$10^5$ colony forming units/ml was produced by adding the solution to sterile Mueller Hinton Broth (MHB, Thermo Fisher). 50 µl was added to each well in the Sensititre plate. The plates were read using the Sensititre manual viewer, with growth recorded as turbidity or as a deposit of cells at the bottom of the well following incubation at 35$^{o}$C $O_2$ for 24 hours. The MIC was interpreted as the lowest concentration of an antimicrobial that inhibits visual growth. All Sensititre plates were assessed for growth control and the inoculum was assessed for purity and colony counts.

#### Genomic DNA extraction:

##### 2017 outbreak:

The initial six suspected carbapenemase-producing *E. coli* isolates were grown on horse blood agar at 37°C overnight. 2 ml of Luria Bertani (LB) media was added to the plate to resuspend the bacterial growth. 300 µl of resuspension was pelleted and used for DNA extraction using the MoBio UltraClean Microbial DNA extraction kit, as per manufacturer's instructions. All other isolates were grown on horse blood agar at 37°C overnight, and DNA was extracted using the DSP DNA Mini Kit on the QIAsymphony SP (Qiagen).

##### 2017-2024 post-outbreak:

DNA extractions for Illumina sequencing were performed as described in Forde *et al* (2023)[1]. Briefly, genomic DNA was extracted using the DSP QIAamp DNA mini kit (Qiagen) on a QiaSymphony SP. For Oxford Nanopore Technology (ONT) sequencing, -80°C glycerol stocks of stored isolates were recovered and plated onto horse blood agar and grown at 37°C overnight. A 10 µl loop of overnight culture was taken directly from each overnight plate and extracted using the Roche High Pure PCR

Template Preparation Kit, as per manufacturer's instructions (LifeScience). All DNA was quantified by fluorometry (Quant-iT dsDNA Assay High Sensitivity Kit for Illumina; Qubit dsDNA Broad Range Kit for ONT; Thermo Fisher Scientific).

#### Illumina sequencing:

##### 2017 outbreak:

The initial six isolates were sequenced at the Australian Centre for Ecogenomics (ACE). All subsequent isolates were sequenced at Queensland Forensic and Scientific Services (QFSS). All libraries were prepared using the Nextera XT DNA preparation kit (Illumina) and sequencing was performed on a NextSeq 500 (Illumina) with 2x150bp chemistry, NextSeq Midoutput kit v2.5.

##### 2017-2024 post-outbreak:

Isolates were sequenced as described in Forde et al (2023)[1]. Briefly, paired-end DNA libraries were prepared using Nextera XT library prep kits (Illumina; Australia) and WGS was preformed using the Illumina NextSeq 500 (150 bp paired-end).

#### Nanopore MinION sequencing:

##### 2017 outbreak:

1.5 µg of DNA for MS14441, MS14442 and MS14443 was used as input for sequencing on an Oxford Nanopore MinION. Libraries were prepared using the 1D sequencing by ligation (SQK-LSK108) kit (without multiplexing) and loaded onto separate FLOW-MIN106 R9.4 flow cells. MS14441 and MS14443 were run for 21 and 23 hours, respectively. MS14442 was only run for 14 hours due to flow cell failure.

##### 2017-2024 post-outbreak:

All post-outbreak isolates were sequenced on a single PromethION R10.4.1 flow cell after library preparation with the rapid barcoding kit (SQK-RBK114.96) as per manufacturer’s instructions.

##### Nanopore analysis during 2017 outbreak:

Nanopore raw reads were basecalled using Albacore (v1.1.1). Reads were filtered using Japsa v1.5-11a[2] to remove reads below Q10 and less than 2000 bp in length.

Filtered Nanopore reads were *de novo* assembled using Canu v1.7[3] at default settings. Assemblies were polished with eight rounds of Pilon (v1.23)[4] using the trimmed Illumina reads. PHASTER[5] (web portal accessed 20/5/2019) was used to detect prophage.

#### Screening public data (AllTheBacteria) for closely related *E. coli*:

All *E. coli* from AllTheBacteria (release 0.2; n=315,066)[6] were sketched using Skani (v0.3.0)[7] at default settings. The chromosome from MS14441 was searched against the *E. coli* sketched database using Skani and filtered for "Align_fraction" >80 and ANI >=99.8 using csvtk (v0.32.0)[8], resulting in 292 genomes. Biosamples uploaded as part of the original outbreak (and present in AllTheBacteria) were filtered, resulting in a final collection of 207 genomes closely related to MS14441. MLST for the 207 genomes was determined using mlst (v2.23.0)[9]. AMR genes were identified using AbritAMR (v1.0.19)[10]. Plasmid replicon types were identified using abricate[11] (v1.0.1) with the plasmidfinder database (2023-11-4)[12]. Parsnp (v2.1.5)[13] was used to generate a whole genome alignment between all 207 genomes (with the MS14441 chromosome as the reference). The alignment was then filtered for recombination using Gubbins (v3.4.3)[14] and a phylogeny generated using RAxML (v8.2.12)[15] with "--bootstrap 100 --seed 456 --model GTRGAMMA". The tree was visualised and annotated using iTol[16].

### Supplementary Results:

#### Initial detection of ESBL and carbapenemase-producing *E. coli*

Over a single week in May 2017, eight patients across three separate locations were identified by rectal swabs performed for routine screening as newly colonised with ESBL-producing *E. coli* displaying an unusual and identical antibiogram. In addition to having an ESBL phenotype, the possibility of carbapenemase activity was also demonstrated, based on meropenem MICs falling above the EUCAST screening breakpoint for carbapenemase-producing Enterobacterales (CPE) (>0.12 mg/L), but below the clinical breakpoint (≤2 mg/L). All strains were found to have phenotypic carbapenemase production by the β-carba test and grew on chromogenic media. The presence of an OXA-48-like carbapenemase gene in all 8 initial isolates was confirmed by real-time multiplex PCR.

A single isolate (MS14442) with increased resistance to meropenem (>32 mg/L by Etest and BMD) differed by 4 core SNPs from the majority (Supplementary Figure 1), including a SNP resulting in a premature stop codon in the outer membrane porin gene *ompF* (Supplementary Table 1). This mutation has previously been associated with increased resistance to carbapenems[17,18]. The same isolate also had a non-synonymous SNP in *ompC* (Supplementary Table 1). While mutations in *ompC* have previously been linked to increased resistance to carbapenems[19], the contribution of this SNP is unknown.

As the antibiogram was noted to be unusual locally (an ESBL *E. coli* susceptible to both gentamicin and trimethoprim-sulfamethoxazole), the laboratory information system was interrogated to identify other potential cases. Of 1061 *E. coli* isolates with an ESBL or CPE phenotype isolated from screening or clinical specimens collected in Queensland public hospitals from 1$^{st}$ January to 15$^{th}$ May 2017, there were only eight with an identical antibiogram. Of these, three were isolated at the PAH in April or May. The earliest isolate (April 6$^{th}$) was from a patient who had been hospitalised in Vietnam immediately prior to returning to Australia for further medical care and had been in hospital from early March. The patient was not a formal inter-hospital transfer and was not screened on admission, and died prior to recognition of the outbreak. No isolates from any of the three retrospectively identified patients were available for further testing.

##### Nanopore sequencing to fully characterize three representative ST38 isolates

All three isolates sequenced with Nanopore MinION (MS14441, MS14442 and MS14443) had operons relating to adhesion and biofilm formation (*csg, ecp,* and *fim*), iron uptake (*chu, ent, fep, sil*) and secretion (including type II [*gsp*], and a putative type VI [*aec*]). Presence of the *E. coli* type III secretion system 2 (ETT2) was confirmed in all three, however the locus of enterocyte effacement (LEE) was absent. All isolates carried the same prophage regions throughout their chromosome except that MS14441 had a single additional ~32 kb phage absent from MS14442 and MS14443 (Supplementary Figure 2). No other genomic rearrangements (large inversions, deletions or insertions) in the chromosome or plasmids were detected.

##### The IncI1 plasmid may have introduced blaCTX-M-15 into the cell

All isolates also carried a ~106 kb IncI1 plasmid that did not carry any genes known to be of clinical significance. However, comparison of this IncI1 plasmid to public data found a closely related plasmid carrying an IS*Ecp1*-$bla_{CTX\text{-}M\text{-}15}$-Tn*3* module (pEC36I; NZ_JAGDML010000002)[20]. The similarity of this plasmid to ours (>98.6% identity over 82% of the length of pEC36I) suggests that this plasmid could represents a relative to our IncI1 plasmid, potentially carrying $bla_{CTX\text{-}M\text{-}15}$ into the cell prior to its chromosomal integration.

##### Additional genetic differences in outbreak dataset:

###### SNPs and structural variants in SS17M6399 and SS17M6415

Isolate SS17M6399 was found to have 264 SNPs within a ~9.6 kb region corresponding to a phage tail protein. On closer inspection, this appeared to be caused by mis-mapping of reads from a similar prophage in SS17M6399. PHASTER analysis identified additional prophage sequences in this genome compared to the reference strain MS14441 (Supplementary Table 2 and Supplementary Figure 2). Isolate SS17M6415 appeared to have a different IncI1 plasmid (albeit with a similar IncI1 plasmid backbone) compared to the majority of outbreak isolates (Supplementary Figure 4).

**Acquisition of different plasmids in three isolates**

Three isolates, SS17M5159, SS17M5157 and SSM6408, all had additional IncQ, IncL/M and IncI2 plasmid types, respectively. Acquisition of the IncQ plasmid in SS17M5159 corresponded to additional resistance genes, including *tetA*, *tetR*, $bla_{TEM\text{-}1b}$, *dfrA5* and *sul2*. These additions are predicted to confer resistance to streptomycin, sulphonamides and tetracycline, consistent with the antibiogram for this isolate. Comparison of this IncQ plasmid to publicly available genomes identified a similar ~115 kb plasmid (pO111-CRL-115, GenBank KC340959.1) isolated in Australia between 1999-2002 (Supplementary Figure 6 and 7).

#### Clinical outcomes

Of the 78 patients confirmed by WGS to be colonised with the outbreak strain, two developed a bloodstream infection (BSI). One patient with co-morbid decompensated cirrhosis and gastro-intestinal bleeding was treated with meropenem and trimethoprim-sulfamethoxazole with a good clinical response. There was no microbiological relapse but subsequently the patient had a clinical decline with sepsis suspected to have a role, and died 30 days after the BSI, which may have contributed to death. This patient had an initial positive rectal swab for the outbreak *E. coli* strain (SS17M5169), which was confirmed using WGS. The blood isolate (M94949) from this patient was sequenced subsequent to the initial outbreak and was found to have 11 additional core SNPs, many of which were non-synonymous and may have been selected for during infection (Supplementary Table 3). The other patient had an indwelling urinary catheter following a spinal injury and developed polymicrobial urosepsis with a carbapenemase-producing *K. pneumoniae* also isolated (both the *E. coli* [SS17M7624-2] and *K. pneumoniae* [SS17M7624-1] were sequenced). The patient was treated with 48 hours of meropenem followed by 14 days of oral trimethoprim-sulfamethoxazole with no recurrence of infection.

#### International source of outbreak isolate

Epidemiological investigations were conducted to identify records of carbapenemase-producing Enterobacterales (CPE) from the preceding years. In the period between 2012 to 2016, there were 31 patients with CPE; 25 patients with a locally endemic IMP-4-producing *Enterobacter cloacae*, 3 patients with OXA-48-like producing *E. coli* and *Klebsiella* colonisation or infection, 3 imported cases of NDM colonization with no secondary transmission, and no isolates with KPC or VIM detected. While these isolates were not available for sequencing, their antibiogram records did not suggest a relationship with the outbreak strain.

Broader matching of antibiograms identified a patient transferred from Vietnam who, on April 6$^{th}$ 2017, was found to be carrying an *E. coli* with an identical antibiogram to the outbreak strain. The patient was not a formal inter-hospital transfer and therefore was not screened on admission. No samples were available for testing as the patient was deceased prior to the recognition of the outbreak. However, comparison of the MS14441 chromosome to the NCBI non-redundant nucleotide database identified a match to the *E. coli* strain Ecol_545 (GenBank CP018976.1), isolated from a patient in Vietnam in 2011. MS14441 and Ecol_545 shared 99.93% average nucleotide identity (ANI) over >95% of the MS14441 chromosome, supporting the idea that this strain may have been introduced to the hospital by the putative index patient who had been recently hospitalised in Vietnam (Supplementary Figure 8).

To provide additional context, we queried the recently constructed AllTheBacteria (release 0.2) dataset for isolates with >99.8% ANI across >80% coverage to our outbreak strain MS14441. From >300,000 *E. coli*, we identified 207 isolates, mainly ST38 but also close variants such as ST3268 and ST3052 (Supplementary Figure 9). Despite restricting to isolates within 99.8% ANI, isolates were globally dispersed across six continents, with limited geographical preference. The isolates that were immediately closest to our strain were (i) found to be from the same outbreak (99.98% ANI, 96.79% alignment fraction) or (ii) isolated in Vietnam (SAMEA5377147; 99.96% ANI, 97.04% alignment fraction) but uploaded publicly after our outbreak, with no collection date available, and without a *bla*$_{OXA-181}$ or an IncX3 replicon. Of 207 isolates, only three carried *bla*$_{OXA-181}$ (and an IncX3/colKP3 replicon), one being from our outbreak. The remaining two isolates (SAMEA6368483, SAMN24021174) were ST38 and ST3268 respectively, with an ANI/alignment of 99.97%/94.87% for SAMEA6368483 and 99.85%/89.82% for SAMN24021174. Both were identified in Europe.

#### No clonal transmission of post-outbreak dataset

Most isolates from the post-outbreak dataset were *Klebsiella pneumoniae* (n=17), followed *by E. coli* (n=9), *Klebsiella grimontii (n=1), Klebsiella variicola (n=1), Citrobacter freundii* (n=1), and *Enterobacter homaechei (n=1).* None of the post-outbreak isolates were the same sequence type as the outbreak clone (*E. coli* ST38) or the *K. pneumoniae* co-colonisation isolate (ST188), however, the majority were identified from the same hospital as the original outbreak (n=14, 47%).

To check for ongoing clonal transmission, Skani was used to rapidly compare our post-outbreak isolates to prospectively collected multidrug resistant bacteria isolated from the same region between 2017-2021[1]. Filtering for ANI > 99.8% and >85% reference alignment identified six clusters containing our post-outbreak samples. Further comparison by applying Split Kmer Analysis (SKA[21] v0.5.1) revealed a pairwise distance of >100 SNPs for most pairs, suggesting no relationship between our samples and other isolates previously observed in the region. We confirmed a direct relationship between Q181_07 and Q181_08 (1 SNP – same patient) and a potential indirect relationship between Q181_20 and Q181_28 (27 SNPs – different patients), and Q181_28 and Q181_30 (43 SNP – different patients).

The two OXA-181-carrying IncAC_2/colKP3 plasmid were found to be genetically identical (0 SNPs, 0 rearrangements). This ~150kb plasmid originated from two different species (*Klebsiella variicola, Escherichia coli*) and two different hospitals (PAH, Royal Brisbane and Women's Hospital [RBWH]) albeit at a similar time (19/11/2023 and 12/11/2023, respectively), suggesting an unknown link.

#### Repeat samples from patients in post-outbreak dataset

Four patients had repeat sampling of different OXA-181-carrying species, suggesting plausible *in vivo* spread of the plasmid as seen during the original outbreak. pQ181_01 and pQ181_11 were two separate IncX3/colKP3 plasmids obtained from isolates Q181_01 and Q181_11 respectively that were collected from patient P03. The plasmids themselves were clonal, thus, presented no structural or SNP differences from each other despite being housed by two separate bacterial hosts - *Enterobacter hormaechei* and *Klebsiella pneumoniae* respectively. This was similarly the case for pQ181_15 and pQ181_16 with their bacterial hosts being *Escherichia coli* and

*Klebsiella pneumoniae* respectively. Other than the IncX3/colKP3 plasmids, there was another set of IncA_C2/colKP3 plasmids pQ181_24 and 25 isolated from the same patient P23 that was clonal despite being collected from separate hospitals a week apart. Additionally, they were also houesd in different bacterial hosts – *Escherichia coli* and *Klebsiella variicola* respectively.

Another cluster of IncX3/colKP3 plasmids (pQ181_14, 15, 16) identified from different isolates (Q181_14, 15, 16) collected from the same patient P10, were all found in different bacterial hosts – *Klebsiella pneumoniae, Escherichia coli* and *Klebsiella grimontii* respectively. Additionally, the truncated Tn*6361* versions identified within the plasmids correlated to their collection dates. Where pQ181_14 had a smaller truncated Tn*6361* (14,142kb Group II in Figure 5) while pQ181_15 and 16 collected two weeks later, had a larger version of Tn*6361* (17,562kb Group III in Figure 5). The larger size of both pQ181_15 and 16 was consequential to their possession of four additional genes, namely: aac(3)-IIe, AAA family ATPase, IS3 family transposase and insE comparatively to pQ181_14.

#### Post-outbreak dataset MIC results

Approximately half of all isolates were reported to be susceptible to meropenem despite being positive for $bla_{\text{OXA-181}}$ (Supplementary Dataset S1; Supplementary Figure 11). Additionally, we found no correlation between the presence of $bla_{\text{CTX-M-15}}$ and increased meropenem resistance, but found that the possession of additional carbapenemase genes, either $bla_{\text{NDM-5}}$ or $bla_{\text{IMP-4}}$, did correlate with higher E-test-derived MICs and were all deemed as resistant to meropenem based on EUCAST clinical breakpoint standards applicable at the time.

#### OXA-181 on the chromosome

When on the chromosome (n=3), the $bla_{\text{OXA-181}}$ gene was found in a Tn2013 transposon (IS*Ecp1* upstream). Isolates Q181_07 and Q181_08 (clonal isolates from the same patient) carried three copies of Tn2013 at different loci on the chromosome, while Q181_12 carried two copies. Q181_07, Q181_08 and Q181_12 also carried blaCTX-M-15 on the chromosome (IS*Ecp1* upstream), with two copies in both Q181_07 and Q181_08, and a single copy in Q181_12.

#### Fusion plasmid carrying both *bla*$_{OXA-181}$ and *bla*$_{CTX-M-15}$

A single plasmid (pQ181_28) appeared twice the size of our regular IncX3/colKP3 plasmid and carried both blaOXA-181 and blaCTX-M-15 with two extra replicons (IncR, IncFIA), suggesting a fusion plasmid. Querying this additional IncR/IncFIA region to PLSDB identified a close match to NZ_CP058943.1 (>99% nucleotide identity and coverage), suggesting a historical fusion event between an IncX3/colKP3 plasmid carrying OXA-181 and a IncR/IncFIA plasmid carrying CTX-M-15 (supplementary figure 12). Only one other plasmid has been reported carrying both OXA-181 and CTX-M-15 (pEC2_1; NZ_CP041956). This plasmid does not appear to be a fusion but instead carries CTX-M-15 on the plasmid downstream of an IS*Ecp1*. Screening of PLSDB for any plasmids containing both blaOXA-181 and blaCTX-M-15 identified pEC2_1 plus three other plasmids ([NZ_CP079627, NZ_CP079632] identical, and NZ_CP103587). Comparison of our fusion plasmid to these plasmids revealed ~99% and ~98% nucleotide identity across roughly ~60% and ~70% of our plasmid (respectively), confirming the novelty of our fusion plasmid.

#### *bla*$_{OXA-181}$ is carried in a truncated variant of Tn*6361*

The *bla*$_{OXA-181}$ gene was found in a Tn*6361*-like transposon on plasmids in our dataset, irrespective of its broader genetic context (n=28) (Supplementary Figure 13). Tn*6361* was originally identified in *Morganella morganii* by Yang *et. al.*[22] in China and is characterised by the presence of *bla*$_{OXA-181}$ and *qnrS1* flanked by IS*26*. The *bla*$_{OXA-181}$ gene is preceded by a truncated IS*Ecp1*, with evidence of a colKP3 plasmid insertion based on the presence of a colKP3 *repA*[23]. Most isolates in our post-outbreak dataset had a “Group II” type truncated Tn*6361* – namely, a Tn*3* family transposase upstream of *bla*$_{OXA-181}$ appears truncated in most of our samples. Several other Tn*6361* variants appeared in our dataset but were represented only in small numbers or as singletons (Supplementary dataset S1).

#### Pling subcommunities outside of outbreak plasmid cluster

The remaining 12 post-outbreak plasmids clustered in 10 other subcommunities (Supplementary Figure 14). Eight subcommunities contained only one plasmid (pQ181_9, 10, 19A, 20, 28-30). One subcommunity contained two post-outbreak plasmids (pQ181_15 and 16). The remaining subcommunity had two post-outbreak plasmids (pQ181_01 and 11) along with two other plasmids isolated from *E. coli* in

China from the environment in 2020 (Refseq: NZ_CP104853.1) and from a human sample in 2022 (Refseq: NZ_CP139362.1). Only NZ_CP104853.1 had a single SNP difference to all the other plasmids in this cluster. Interestingly, pQ181_01 and pQ181_11 were not isolated from *E. coli*, but from *Enterobacter hormaechei* and *Klebsiella pneumoniae* respectively.

### Supplementary Tables:

Supplementary Table 1: Predicted consequences of SNPs in Supplementary Figure 1

| Isolate | SNP location (bp) relative to MS14445[a] reference | Description |
|---|---|---|
| **MS14442** | 615162 | Results in stop codon (TAA) in gene for OmpF outer membrane porin (linked with carbapenem resistance) |
| | 987790 | D (GAC) -> Y (TAC) in gene for OmpC |
| | 989792 | P (CCT) -> S (TCT) in gene for Phosphotransferase RcsD |
| | 2716928 (plasmid) | Single SNP upstream of $bla_{OXA-181}$ (GGGGACGTTATG -> GGGGGCTTATG) |
| **MS14445** | 2716928 | G (GGC) -> G (GGT) in gene for putative FAD-linked oxidoreductase |

[a] Original concatenated *de novo* Illumina assembly of *E. coli* ST38 strain MS14445

Supplementary Table 2: PHASTER comparison of phage sequences in SS17M6399 compared with draft Nanopore reference strain MS14441

| Phage | SS17M6399 | MS14441 | pMS14441A | pMS14441B |
|---|---|---|---|---|
| Entero-mEp460_NC_019716(10) | 49.16%/ 43.94% | 47.35% | | |
| Vibro_12B12_NC_021070(27) | | 52.42% | | |
| Escher_pro483_NC_028943(35) | 45.69% | 51.02% | | |
| Enter_933W_NC_000924(2) | 47.48% | 47.05% | | |
| Shigel_Stx_NC_029120(5) | 47.33% | | | |
| Entero_lambda_NC_001416(20) | 50.83% | | | |
| Entero_N15_NC_001901(2) | 51.68% | | 56.56% | |
| Stx2_c_1717_NC_001357(2) | 43.33% | | | |
| Cronob_vB_CsaM_CAP32_NC_019401(1) | 48.50% | | | |
| Klebsi_phiKO2_NC_005857(2) | | | 54.03% | |
| Staphy_SPbeta_like_NC_029119 (2) | | | | 49.74%/ 50.11% |

Supplementary Table 3: SNP consequences in blood isolate M94949

| **SNP location (in MS14445 reference)** | **Type** | **Description** |
|---|---|---|
| 78591 | Non-synonymous | Non-synonymous SNP in *prc* gene (involved In cleavage of C-terminus of penicillin-binding protein 3 (PBP3) |
| 295377 | Synonymous | Within *uidABC* system, involved in transport of glucuronides |
| 1360125 | Non-synonymous | In *folA* gene, involved in folate metabolism |
| 1641057 | Non-synonymous | In *puuA* gene, involved in utilization of putrescine as carbon and nitrogen source |
| 2134299 | Non-synonymous | Uncharacterised protein YqeB |
| 2192206 | Non-synonymous | In *epd* gene, catalyses NAD-dependent conversion of D-erythrose 4-phosphate to 4-phosphoerythronate |
| 3372972 | Synonymous | In DEAD-box RNA helicase gene involved in RNA degradation |
| 3658694 | Non-synonymous | Required for induction of expression of the formate dehydrogenase H and hydrogenase-3 structural genes |
| 4546526 | Intergenic | Does not appear to be within promoter region of surrounding genes |
| 4678640 | Synonymous | In *fdnG* gene, enables *E. coli* to use formate as major electron donor during anaerobic respiration |
| 4679448 | Non-synonymous | |

#### Supplementary Figures:

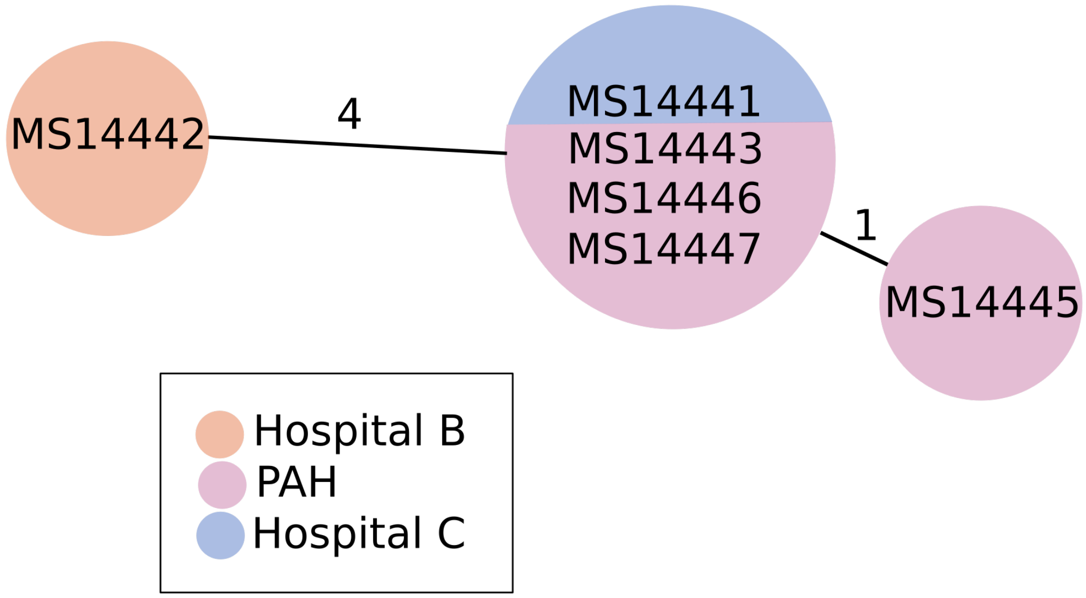


**Supplementary Figure 1: Relationship matrix of initial 6 *E. coli* isolates**
Numbers indicate core SNP distances between isolate genomes. Isolate genomes sharing the same circle are identical at the core SNP level.

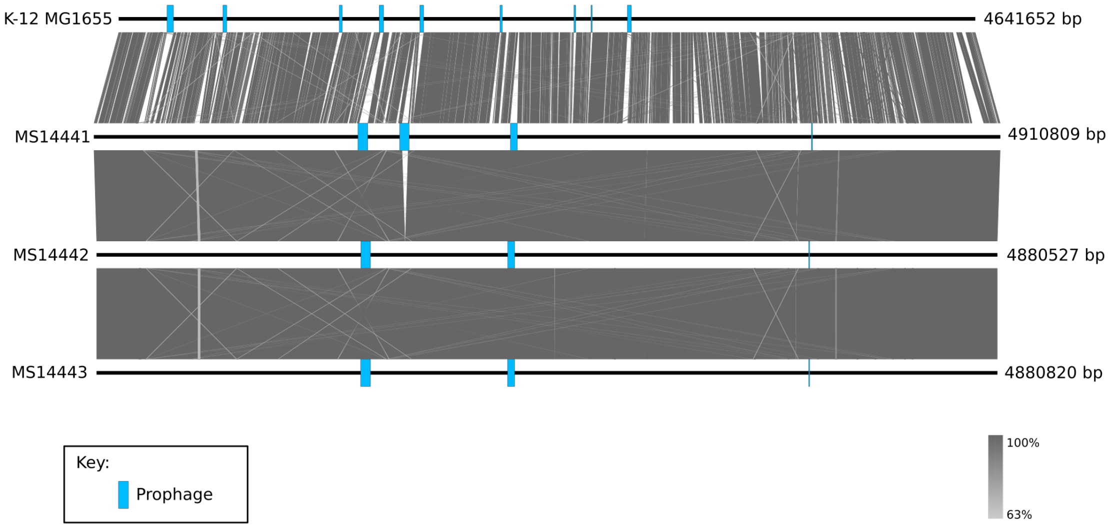


**Supplementary Figure 2: Comparison of entire chromosomes of K-12 MG1655, MS14441, MS14442 and MS14443**

Comparison was carried out with BLASTn as implemented in Easyfig. Prophage regions are annotated in blue.

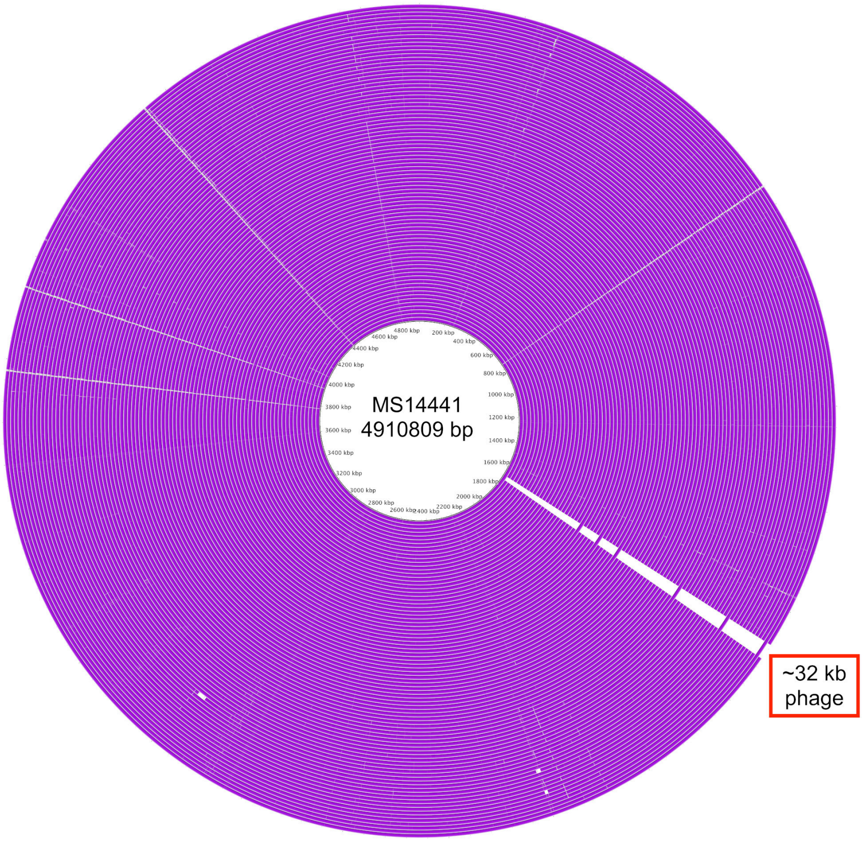


**Supplementary Figure 3: BRIG comparison of all ST38 outbreak isolates to MS14441 chromosome**

For the list of strains for each ring, see Supplementary Dataset S1 (tab 4).

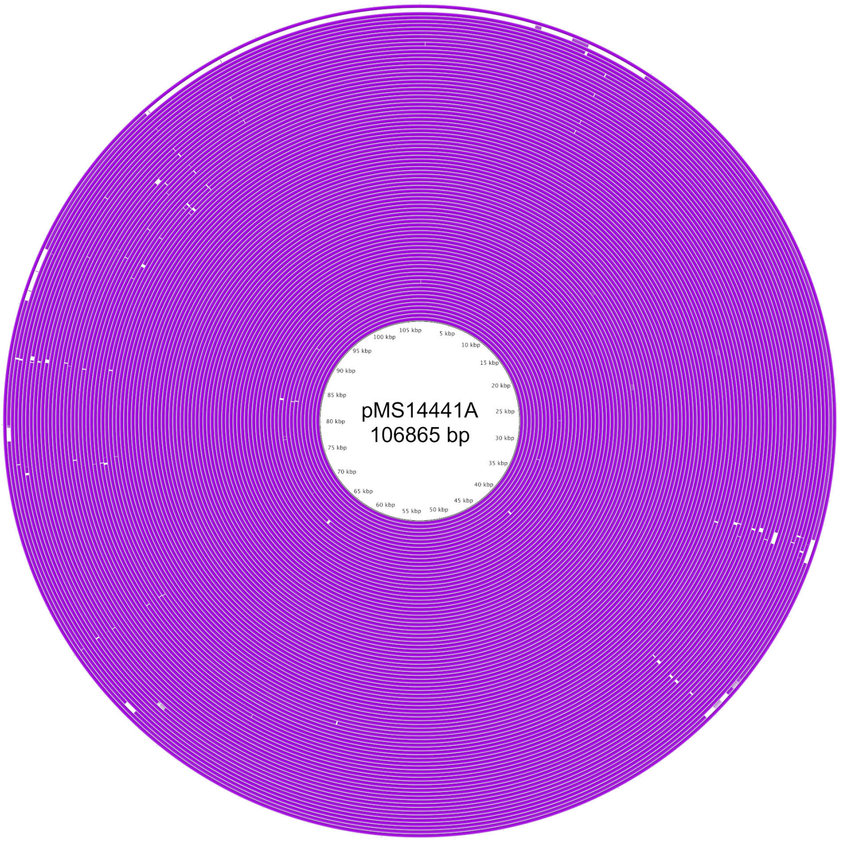


**Supplementary Figure 4: BRIG comparison of all ST38 outbreak isolates to pMS14441A (IncI1 plasmid)**

All isolates appeared to have a very similar/identical IncI1 plasmid, with the exception of SS17M6415 (second outermost ring), which appears to have a different IncI plasmid, albeit with a similar IncI backbone. For the list of strains for each ring, see Supplementary Dataset S1 (tab 4).

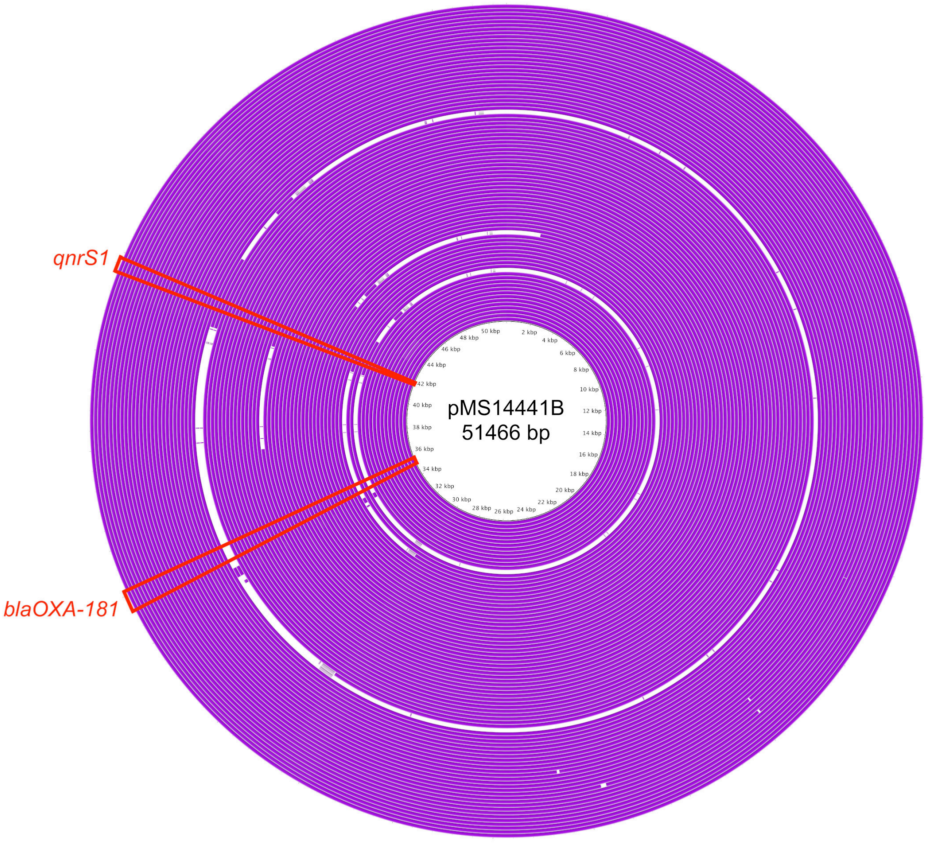


**Supplementary Figure 5: BRIG comparison of all ST38 outbreak isolates to pMS14441B (IncX3 plasmid)**
For the list of strains for each ring, see Supplementary Dataset S1 (tab 4).

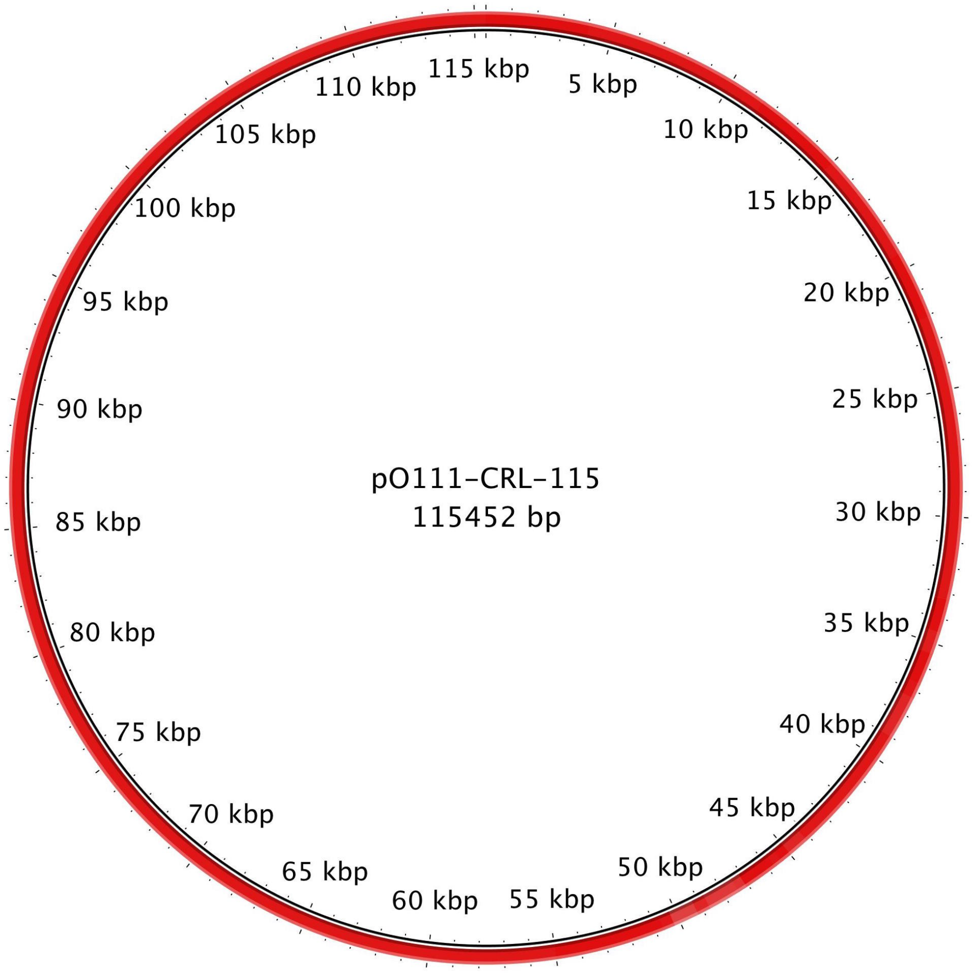


**Supplementary Figure 6: BRIG comparison of SS17M5159 draft assembly to pO111-CRL-115**

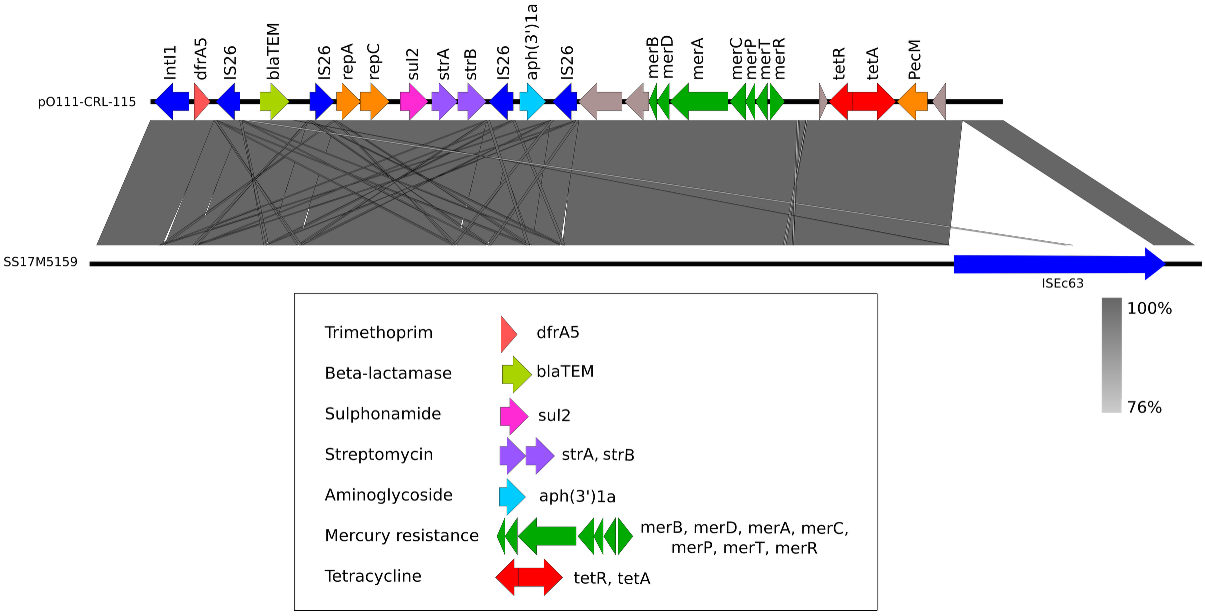


**Supplementary Figure 7: Context of antibiotic resistance genes in SS17M5159 based on reference plasmid pO111-CRL-115**

BLASTn comparison of pO111-CRL-115 resistance region (top) in comparison with matching plasmid contigs from draft SS17M5159 genome (bottom) prepared using Easyfig.

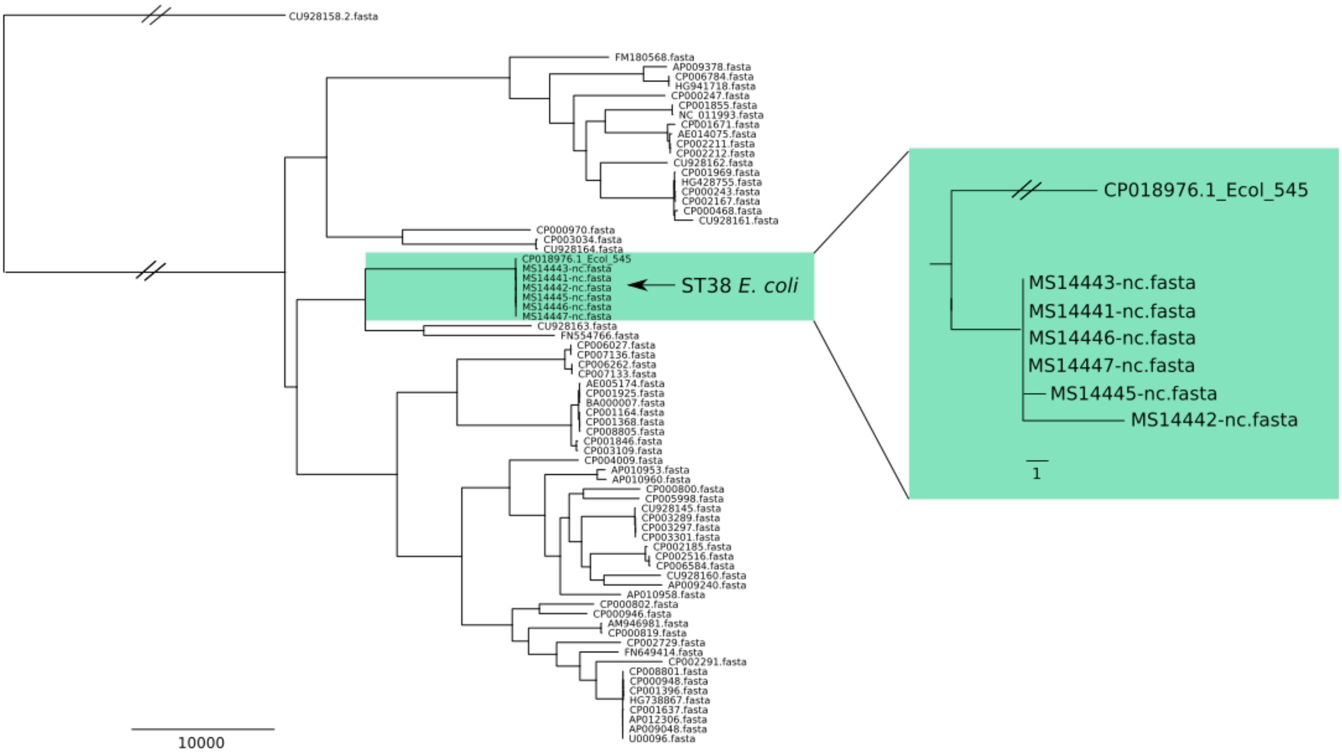


**Supplementary Figure 8: Contextualizing ST38 outbreak isolates against publicly available E. coli complete genomes**

The MS14441 reference chromosome (generated using Nanopore MinION) was used to search the non-redundant nucleotide (nr/nt) database available on NCBI's using BLAST [https://blast.ncbi.nlm.nih.gov/Blast.cgi] (accessed 28$^{th}$ May 2017). *E. coli* 545 (CP018976.1) appeared as the closest match. A selection of available complete *E. coli* genomes were used to contextualize the outbreak isolates.

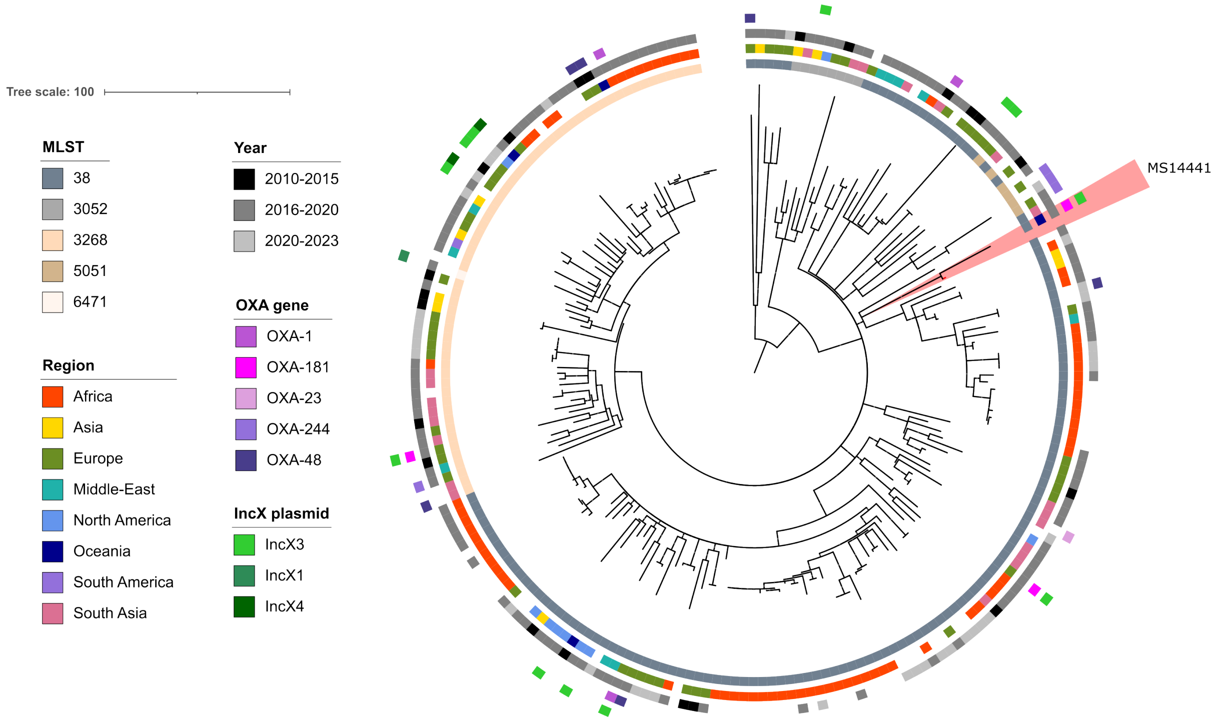


**Supplementary Figure 9: RAxML tree of 207 genomes from AllTheBacteria**

>99.8% ANI across >80% of the MS14441 chromosome.

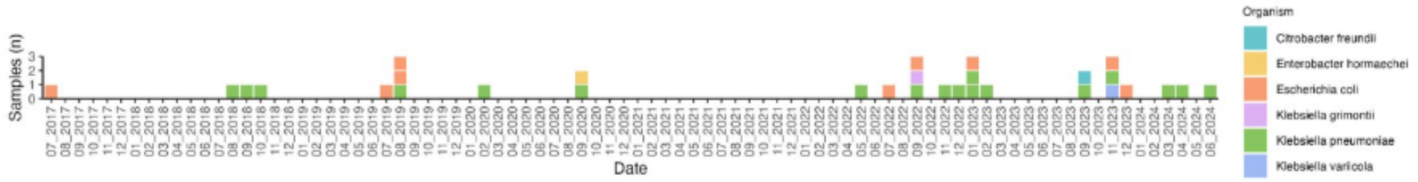


**Supplementary Figure 10: timeline of post-outbreak dataset**

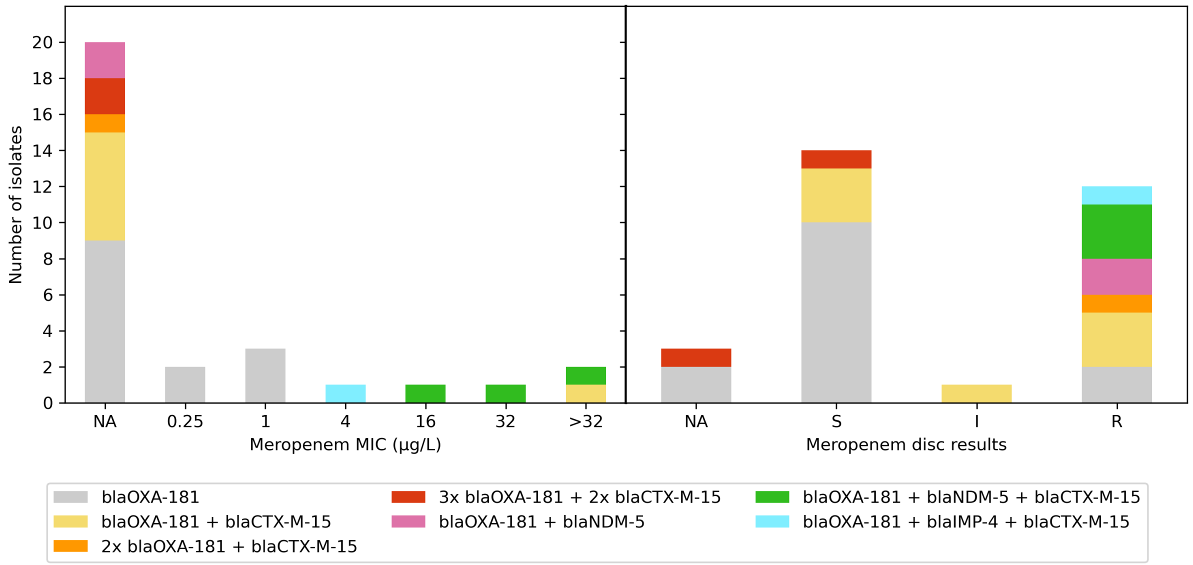


**Supplementary Figure 11: Available Meropenem Minimum Inhibitory Concentration (MIC) and Meropenem disc diffusion susceptibility testing results for post-outbreak isolates (n=30) collected from Pathology Queensland.** The number of isolates tested was plotted against MIC values for the meropenem Epsilometer test (E-test), with results ranging from 0.25 to >32μg/L concentrations for minimum inhibition (left). Whilst for the meropenem disc results, the number of isolates deemed to be either susceptible (S), susceptible, increased exposure (I) or resistant (R) by EUCAST guidelines was plotted (right) against each isolate. NA represents the number of isolates not tested for the respective tests on both sides of the figure. Long-read sequencing allowed for the identification of multiple copies of *blaOXA-181*, additional carbepenemase genes (*blaNDM-5* and *blaIMP-4*) along with cephalosporin resistance gene *blaCTX-M-15*. The combinations found were as follows and were colour coded in the stacked bar graphs: one copy of OXA-181 (grey), one copy of *blaOXA-181* and one copy of *blaCTX-M-15* (yellow), two copies of *blaOXA-181* and one copy of *blaCTX-M-15* (orange), three copies of *blaOXA-181* and two copies of *blaCTX-M-15* (red), one copy of *blaOXA-181* and one copy of *blaNDM-5* (pink), one copy of *blaOXA-181*, one copy of *blaNDM-5* and one copy of *blaCTX-M-15* (green), lastly one copy of *blaOXA-181*, one copy of *blaIMP-4* and one copy of *blaCTX-M-15* (blue).

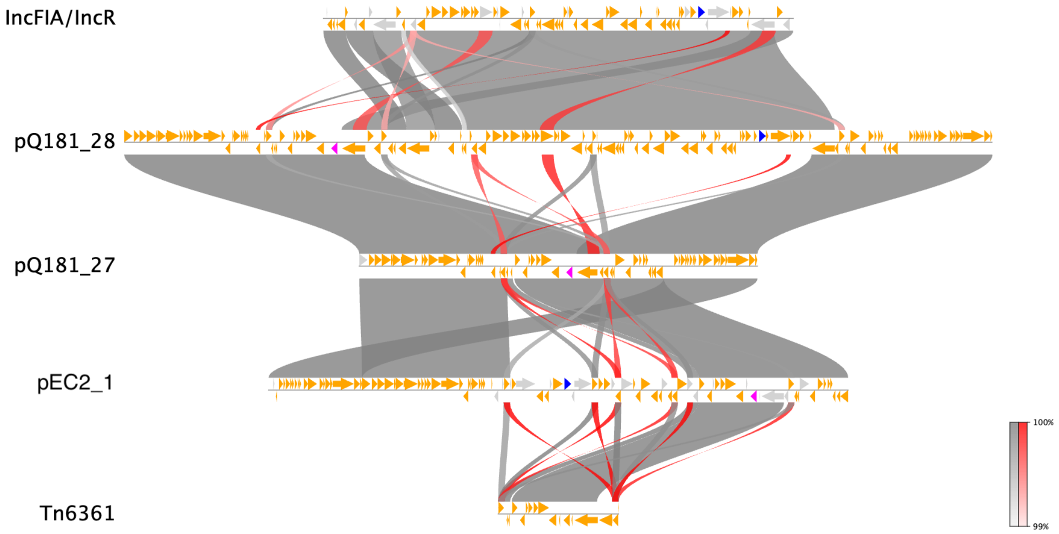


**Supplementary Figure 12: Fusion plasmid containing *bla*$_{OXA-181}$ and *bla*$_{CTX-M-15}$**

pQ181_28 was identified to carry both blaOXA-181 and blaCTX-M-15 on the same plasmid, with replicons IncX3, colKP3, IncR and IncFIA. Comparison to other plasmids (pQ181_27) revealed a single, large insertion containing the IncR/IncFIA replicons. This region corresponded to NZ_CP058943.1 [IncFIA/IncR] (>99% nucleotide identity and coverage), suggesting a historical fusion event between an IncX3/colKP3 plasmid carrying OXA-181 and a IncR/IncFIA plasmid carrying CTX-M-15. Only one other plasmid has been reported carrying both OXA-181 and CTX-M-15 (pEC2_1; NZ_CP041956). This plasmid does not appear to be a fusion but instead carries blaCTX-M-15 on the plasmid downstream of an IS*Ecp1*.

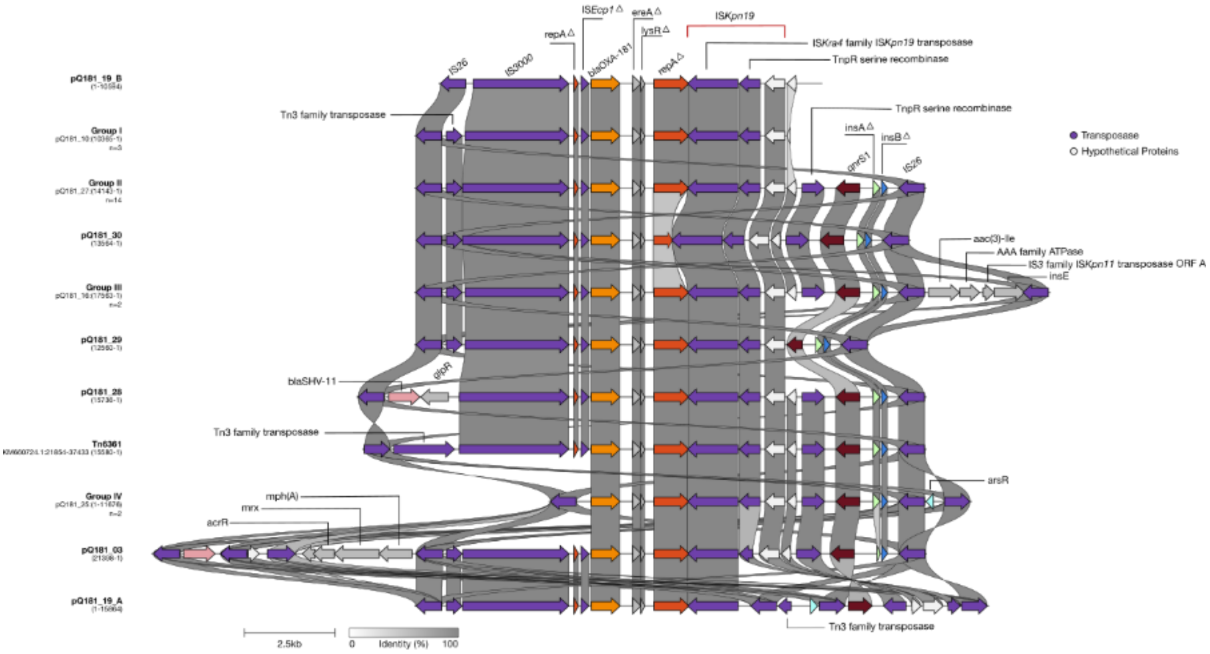


**Supplementary Figure 13: Structural comparison of post-outbreak plasmids with the blaOXA-181 carrying transposon Tn6361.** Truncated versions of Tn*6361* were identified by alignment with each of the post-outbreak plasmids containing *bla*$_{OXA-181}$. Plasmids containing the same structural arrangement were grouped into Groups I-IV with the representative isolate labelled. Coordinates of the Tn*6361*-like region are labelled below each representative isolate. Grey bars represent amino acid percent identity of gene features (intergenic relatedness not shown). Figure generated using Clinker (v0.0.32).

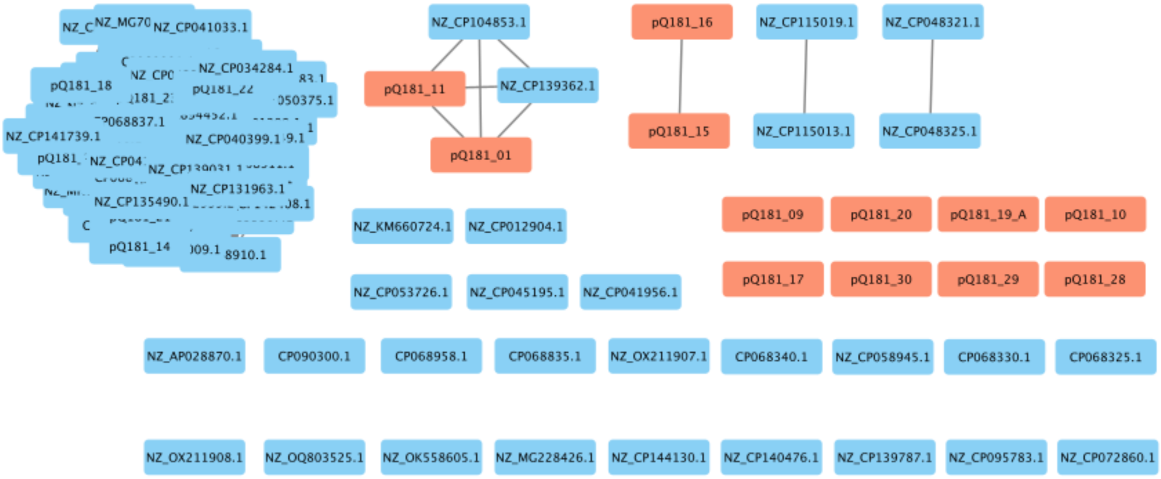


**Supplementary Figure 14: Pling network of the 114 IncX3/colKP3 plasmids obtained from PLSDB along with post-outbreak plasmids and original outbreak plasmid clustering at DCJ 0.** Post-outbreak plasmids that did not cluster with the largest 98 plasmid cluster are colored orange.
